# Insulin moderates the effects of early life adversity on executive functioning in a sex-specific manner

**DOI:** 10.1101/2024.10.08.24315109

**Authors:** Aashita Batra, Irina Pokhvisneva, Guillaume Elgbeili, Olivia Ruge, Eamon Fitzgerald, Sachin Patel, Darina Czamara, Michael J Meaney, Elisabeth B. Binder, Patricia P Silveira

**Author notes:** Correspondence: Patrícia Pelufo Silveira, MD, PhD, Department of Psychiatry, Faculty of Medicine and Health Sciences, McGill University. Douglas Hospital Research Centre, 6875 Boulevard LaSalle, Montreal, QC, H4H 1R3, Canada. (ext.2776).

## Abstract

**Background:** Early life adversity (ELA) is associated with altered insulin signaling and altered EF behaviors, in a potentially sex-specific manner. Considering the high co-morbidity between altered metabolism and executive function (EF) problems, we hypothesized that the genetic background associated with altered fasting insulin (FI) and EF could be shared

**Methods:** Our study used conjunctional false discovery rate (ConjFDR) to identify the shared genetic architecture between FI and two EFs: impulsivity and attention deficit-hyperactivity disorder (ADHD). We identified the polygenic risk score (PRS) threshold from a FI genome-wide association study (GWAS) that best predicted insulin levels in male and female ALSPAC children [N_males_=1,901, N_females_=1,834; p_t-intial-males_= 0.05 (11,121 SNPs), p_t-intial-females_= 0.15 (27,202 SNPs)], further refining it to only include SNPs significantly associated with insulin levels in children [N_SNP-males_= 635 SNPs, N_SNP-females_ = 1,449 SNPs]. A phenome-wide association study (PheWAS) was also run to identify EFs associated with the interaction between the refined PRS (rPRS) and early adversity. To investigate the presence of a direct causal relationship between FI and impulsivity in the presence of adversity, we applied mendelian randomization (MR)

**Results:** ConjFDR suggested that environmental factors could be involved in the association between insulin and EFs, as there was no shared genetic background. PheWAS highlighted impulsivity and attention-related outcomes in interaction models between FI rPRS and early adversity. Finally, two-sample MR suggested a causal association between higher fasting insulin levels and impulsive behavior, specifically in females exposed to adversity (p < 0.001). Overall, a sex-specific impulsivity GWAS demonstrated that *MYT1L* and *TSSC1*, genes that are associated with motor impulsivity, were enriched only in females.

**Conclusions:** Our study solidifies the evidence that the relationship between high FI and EF is not direct, but rather interacting with ELA exposure, especially in females.

**Key points:**

- Early life adversity is associated with alterations in insulin signaling and executive functioning behaviors.
- We report a causal association between high fasting insulin and increased impulsivity in females exposed to adversity.
- Our findings also support the idea that fasting insulin moderates the long-term effects of early life adversity on executive functions in females.
- This research provides insights into the mechanisms by which insulin moderates the effects of early life adversity on executive function disorders and informs the development of potential interventions.

## Introduction

Exposure to ELA is a developmental risk factor associated with metabolic and psychiatric phenotypes. ELA leads to alterations in responsivity to stress and alters insulin sensitivity at different ages. Childhood adversity and adult stress exposures have been linked to diabetes and altered glucose regulation, and chronic adulthood stress has been associated with glucose intolerance and metabolic syndrome (Pyykkönen *et al*, 2010). These findings emphasize that ELA impacts insulin regulation and function, making it an important node to investigate.

Insulin is a hormone involved in metabolism regulation in animals, and a neuroregulatory peptide in the central nervous system, including the prefrontal cortex (PFC) (Ghasemi *et al*, 2013). In PFC, insulin modulates the development and expression of different executive function (EF) behaviors (Kullmann *et al*, 2016), such as attention, inhibitory control, and working memory. Insulin reduces activity in the prefrontal areas that control behaviors like the inhibitory control of eating (Heni *et al*, 2015). In Alzheimer’s patients, insulin dysregulation has been linked to learning deficits and memory formation impairments (Zhao and Alkon, 2001).

A recent study demonstrated that metabolic syndrome and IR were associated with poorer executive performance in women (Schuur *et al*, 2010), suggesting that insulin could be related to EF in a sex-specific manner. Co-morbidity between metabolic and psychiatric diseases has been established, and, interestingly, among adults with ADHD, there is a 18.7% higher prevalence of type 2 diabetes in males compared to females (Chen *et al*, 2018).

We have previously reported (Batra *et al*, 2021) that a polygenic risk score associated with higher FI interacts with ELA to predict the development of inhibitory control in children at 36 months. This reinforced the idea that insulin may moderate the effects of ELA on EF. However, as the effects of ELA are often sex-specific (Barbieri *et al*, 2009b; Honeycutt *et al*, 2020), a better understanding of sex-specific relationships between ELA, insulin, and the development of EF is needed. Our hypotheses for the current study are 1) the genetic backgrounds associated with high FI and EF share a polygenic architecture; 2) if (1) is rejected, models including the interaction term between the genetic background associated with high FI and childhood adversity exposure could show association with EF across lifespan; 3) if (2) shows significance results, there may be a causal relationship between FI levels and impulsivity in the presence of adversity, which we will inspect through mendelian randomization (MR).

## Methods

### Investigating the shared genetic background between higher fasting insulin and altered executive function phenotypes (impulsivity and ADHD)

#### GWASs

We obtained complete FI GWAS results in the form of summary statistics p-value from the Meta-Analyses of Glucose and Insulin-related traits Consortium (Lagou *et al*, 2021) (*Refer to Supplementals*). ADHD GWAS was obtained from sex-specific meta-analyses of case-control ADHD by the Psychiatric Genomics Consortium and the Lundbeck Foundation Initiative for Integrative Psychiatric Research (Kettunen *et al*, 2016) (*Refer to Supplementals)* As the sex-specific impulsivity GWAS was not available in the literature, we performed this GWAS using the UK Biobank data (*Refer to Supplementals)*.

#### Conditional/conjunctional FDR

Analyses applied here used false discovery rate (FDR) methods previously established by Andreassen et al (Andreassen *et al*, 2013b). To evaluate the cross-phenotype polygenic architecture, we produced conditional quantile-quantile (Q-Q) plots. The plots condition FI on impulsivity or ADHD, and vice versa, plotting observed p-values against theoretical quantiles under no association. A straight null line indicates no association, whereas deviation indicates systematic association. Each Q-Q plot displays SNP p-values of trait 1, conditional on varying associations with trait 2, which allows us to determine a presence of a shared polygenic architecture between the traits.

To identify shared loci between FI and impulsivity or ADHD, we applied the conditional FDR (condFDR) and conjunctional FDR (conjFDR) methods (Andreassen *et al*, 2013a; Andreassen *et al*, 2014) (Note: both methods assume the p-values for that SNP in both the primary and the conditional traits are lower than the observed p-values). CondFDR employs genetic association summary statistics from a trait of interest (FI) together with those of a conditional trait (impulsivity or ADHD) to estimate the posterior probability that a SNP has no association with the primary trait. This method identifies loci associated with the primary trait by using associations with the conditional traits. The conjFDR statistic is the maximum of the 2 mutual condFDR values and is a conservative estimate of the posterior probability that a SNP has no association with either trait. Using conjFDR helps identify loci which are associated with both traits. A conservative FDR level of 0.01 per pairwise comparison was set for condFDR/conjFDR, corresponding to a false positive rate of 1 in 100 reported associations. More details can be found in the original and subsequent publications (Andreassen *et al*, 2013a; Andreassen *et al*, 2014; Andreassen *et al*, 2013b; O’Connell *et al*, 2020).

### Investigating the interactive effects between polygenic scores for higher fasting insulin and childhood adversity on altered executive function phenotypes in a sex-specific manner

#### Participants

We used data from five prospective birth cohorts: 1) Avon Longitudinal Study of Parents and Children (ALSPAC) (Boyd *et al*, 2013; Fraser *et al*, 2013); 2) Maternal Adversity, Vulnerability, and Neurodevelopment (MAVAN) (O’Donnell *et al*, 2014); 3) Growing Up in Singapore Towards healthy Outcomes (GUSTO) (Soh *et al*, 2014); 4) Adolescent Brain Cognitive Development ^SM^ Study (ABCD®) (Jernigan *et al*, 2018); and 5) UK Biobank (UKB) (Sudlow *et al*, 2015) to analyze the gene by environment interaction effects on EF outcomes. Details about recruitment, inclusion criteria for this study and genotyping information on the cohorts can be found in supplementary materials.

*The Avon Longitudinal Study of Parents and Children (ALSPAC)* (Northstone *et al*, 2019): For our analysis, we included children of 8.5 and 9.5 years old (individuals with peripheral insulin levels data), whose mothers had a pregnancy duration between 37 and 42 weeks, a maternal age at delivery greater than 18 years, a child birthweight greater than 2 kg, child alive at 1 year of age. We only included singleton pregnancies in the analysis. There were 1,901 males and 1,834 females with complete data available for the analyses. Ethics approval for the study was obtained from the ALSPAC Ethics and Law Committee and the local research ethics committees (a full list of the ethics committees that approved different aspects of the ALSPAC studies is available at http://www.bristol.ac.uk/alspac/researchers/research-ethics/). Data were collected during clinic visits or with postal questionnaires. Informed consent for the use of data collected via questionnaires and clinics was obtained from participants following the recommendations of the ALSPAC Ethics and Law Committee at the time.

*Maternal Adversity, Vulnerability, and Neurodevelopment (MAVAN)* Project (O’Donnell *et al*, 2014): After applying the exclusion criteria, 161 subjects had complete data on the relevant predictors, as described in *supplementary Figure S7*. Approval for the MAVAN project was obtained by the ethics committees and university affiliates (McGill University and Université de Montréal, the Royal Victoria Hospital, Jewish General Hospital, Centre hospitalier de l’Université de Montréal and Hôpital Maisonneuve-Rosemount) and St. Joseph’s Hospital and McMaster University, Hamilton, Ontario, Canada. Informed consent was obtained from all participants.

*Growing Up in Singapore Towards healthy Outcomes (GUSTO)* (Soh *et al*, 2014) prospective cohort: Study sample was selected based on data availability for each analysis. There were 466 subjects with complete data on the predictors used within this study available for the analyses, as described in *supplementary Figure S7*. This study was approved by the National University Hospital and KK Women’s and Children’s Hospital, National Healthcare Group Domain Specific Review Board (NHG DSRB Ref D/09/021) and Sing Health Centralized Institutional Review Board (CIRB Ref 2018/2767). Written informed consent was obtained from mothers and their children.

*Adolescent Brain Cognitive Development*^SM^ Study *(ABCD®) (Jernigan et al, 2018)*: There were 7,655 subjects with complete data available for the analyses, as described in *supplementary Figure S7*. The institutional review board of the University of California, San Diego centralized the approval for data collection. The study sites where data were collected also obtained approval from their local institutional review boards. All parents gave written informed consent and children verbal assent.

*UK Biobank (UKB)* (Sudlow *et al*, 2015): Of the total 502,543 participants in this cohort, 71,036 subjects had complete data on the predictors used within this study available for the analyses, as described in *supplementary Figure S7*. This research was conducted using the UK Biobank Resource under Application Number 41975. Approval for the UK Biobank was obtained by the North West Multicentre Research 580 Ethics Committee (REC reference 11/NW/0382; http://www.ukbiobank.ac.uk/ethics/), the National Information Governance Board for Health and Social Care and the Community Health Index Advisory Group. Informed consent was obtained from all participants.

#### Refined Polygenic Risk Scores (rPRS)

We sought to identify the optimal high FI PRS threshold to predict FI levels in children from the ALSPAC cohort, in a sex-specific manner (using a sex-specific FI GWAS). We used the refined-PRS (rPRS) method described in our previous study (Batra *et al*, 2021). The rPRS was subsequently applied in four separate cohorts, analyzing males and females seperately, to examine how the interaction between higher FI rPRS and ELA impacts EF behaviors across different age groups.

*ALSPAC:* FI PRSs were calculated using FI GWASs separately for males and females (N_males_ = 47,806, N_females_ = 50,404) from the Meta-Analyses of Glucose and Insulin-related traits Consortium PRSs were calculated at 100 different p-value thresholds for each individual in the ALSPAC cohort as a sum of the risk alleles count, weighted by the effect size described in the GWAS for each SNP. We then utilized Generalized Estimating Equations (GEE) analysis to identify the threshold of PRS at which the model predicting peripheral insulin levels in children at age 8.5-9.5 years best fit to the data, separately in males and females. To further refine the PRS, a process explained in Batra et al (Batra *et al*, 2021), we ran a GEE analysis for each SNP within the identified PRS threshold to identify which SNPs were significantly associated with peripheral insulin levels separately for males and females [N_SNP males_= 635 SNPs, N_SNP females_ = 1,449 SNPs] *(Refer to Supplementals)*.

*MAVAN, GUSTO, ABCD, and UKB:* The SNPs we discovered to associate with peripheral insulin levels in the ALSPAC cohort were used to construct a rPRS in these four cohorts. Because the SNPs were selected in a refinement process of a PRS that was created through conventional means, we refer to this PRS as the refined PRS. The rPRS was calculated similarly to the PRS scores in ALSPAC, as a weighted sum of 635 SNPs for males and 1,449 SNPs for females.

#### Early Life Adversity

To inspect the interaction effect between the PRS and early life adversity, we calculated adversity exposure using a cumulative score involving different environmental variables for each individual in the cohorts as described by Silveira et al. (Silveira *et al*, 2017). Details on the components of the scores can be found in supplementary materials.

#### Executive Function Behavioral Outcomes

To inspect the interaction effect of the PRS and early life adversity on EF, we used all the EF behaviors with available data for each cohort. The MAVAN cohort outcomes consisted of EF behavior data from the Bayley Scales of Infant Development II (Balasundaram and Avulakunta, 2021), Child Behavior Checklist (CBCL) (Achenbach and Edelbrock, 1991), School Readiness Test Battery (Gan *et al*, 2016), Dominique Questionnaire (Kuijpers *et al*, 2013), Strengths and Difficulties Questionnaire (SDQ) (Bøe *et al*, 2016), Cambridge Neuropsychological Test Automated Battery (CANTAB) Intra and Extradimentional shift (IED), and Information Sampling Task (IST) (Sahakian and Owen, 1992). The GUSTO cohort outcomes consisted of EF behaviors data from Bayley Scales of Infant Development II (Balasundaram *et al*, 2021), Child Behavior Checklist (CBCL) (Achenbach *et al*, 1991), Stop Signal Task (SST) (Chikazoe *et al*, 2009), Snack Delay Task (Silveira *et al*, 2012), Sticker Delay Task (Silveira *et al*, 2018). The ABCD cohort outcomes consisted of EF behaviors data from the Child Behavior Checklist (Achenbach *et al*, 1991), Brief Problem Monitor (BPM) assessment (Achenbach *et al*, 2011), and UPPS-P Impulsive Behavior Scale (Cyders, 2013). The ABCD Study data repository grows and changes over time. The ABCD Study data used in this report came from https://dx.doi.org/10.15154/1503209. The UKB cohort analysis consisted of psychiatric outcomes related to EF outcomes.

#### Interaction PheWAS

We performed a linear regression (for continuous outcomes) and logistic regression (for binary outcomes) analyses for each PheWAS to investigate the interaction effect between the rPRS for FI and postnatal adversity exposure on each EF behavior separately in males and females in MAVAN, GUSTO, ABCD, and UKB, adjusted by population stratification PCs, age and other covariates if they were found to be significantly different between low/high PRS groups (as presented in *Tables 1–4*). We also used GEE if an outcome was measured several times. Details on the EF behaviors analyzed for each cohort can be found in supplementary materials.

**Table 1.** Participants’ characteristics in MAVAN. Numbers are presented as mean (SD) or percentage (number of participants). Comparison between Low/High PRS groups were carried out using Student t-test for continuous variables and chi-square test for categorical variables.

| Sample descriptive | Males |  |  |  | Females |  |  |  |
| --- | --- | --- | --- | --- | --- | --- | --- | --- |
|  | Total<br>(n =<br>80) | Low<br>PRS<br>(n =<br>40) | High<br>PRS<br>(n = 40) | *p* | Total<br>(n = 81) | Low<br>PRS<br>(n = 40) | High<br>PRS<br>(n = 41) | *p* |
| Age at delivery<br>(years) | 30.58<br>(4.31) | 30.54<br>(3.95) | 30.61<br>(4.7) | 0.942 | 30.88<br>(4.82) | 31.14<br>(4.74) | 30.62<br>(4.93) | 0.625 |
| Birth weight<br>(grams) | 3449<br>(459) | 3341<br>(373) | 3558<br>(514) | **0.034** | 3233<br>(429) | 3169<br>(406) | 3294<br>(446) | 0.191 |
| Breastfeeding at 3<br>months- yes | 76.9%<br>(60) | 79.5%<br>(31) | 74.4%<br>(29) | 0.788 | 68.4%<br>(54) | 68.4%<br>(26) | 68.3%<br>(28) | 1 |
| Smoking during<br>pregnancy | 12.5%<br>(10) | 12.5%<br>(5) | 12.5%<br>(5) | 1 | 11.1%<br>(9) | 5% (2) | 17.1%<br>(7) | 0.169 |
| Household income <<br>\$30000 | 0.14<br>(0.34) | 0.08<br>(0.27) | 0.19<br>(0.4) | 0.155 | 0.13<br>(0.34) | 0.08<br>(0.27) | 0.18<br>(0.39) | 0.18 |
| Gestational age at<br>birth (weeks) | 39.35<br>(1.21) | 39.27<br>(1.2) | 39.42<br>(1.24) | 0.583 | 39.12<br>(1.18) | 38.92<br>(1.14) | 39.32<br>(1.19) | 0.134 |

**Table 2.**
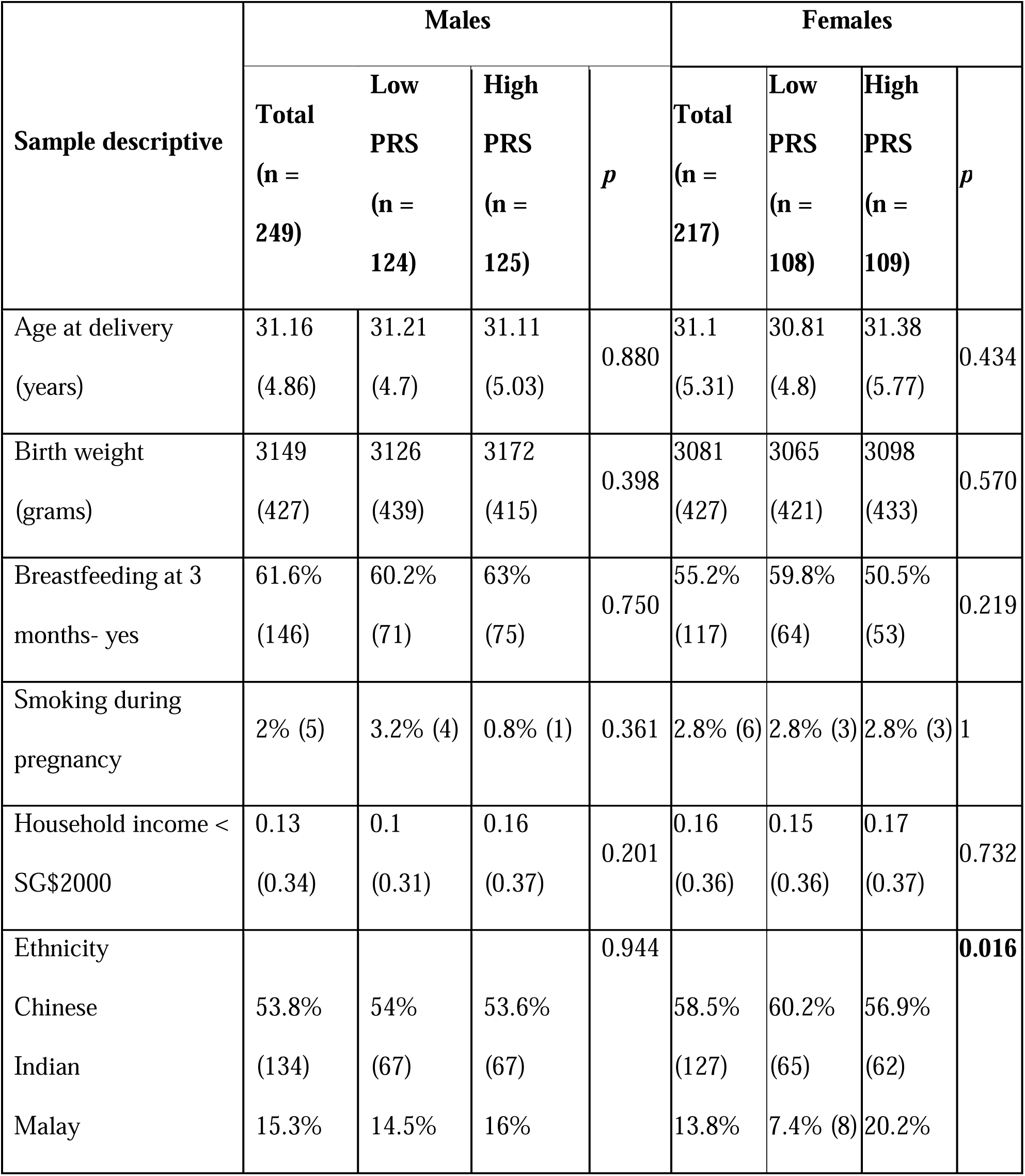
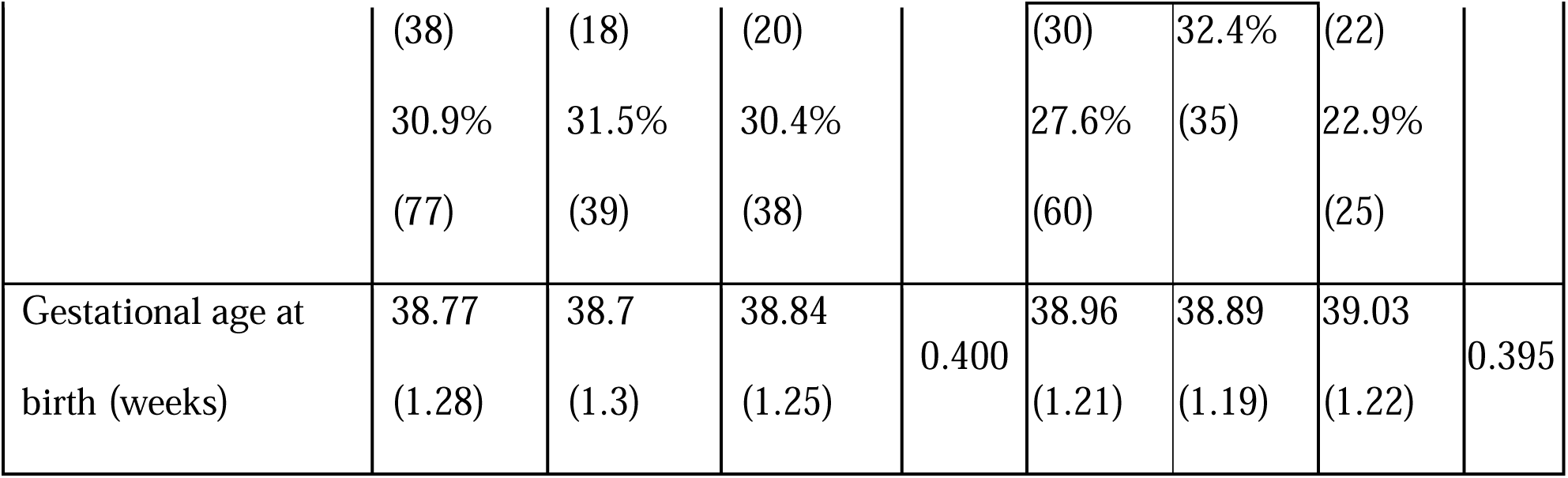
Participants’ characteristics in GUSTO. Numbers are presented as mean (SD) or percentage (number of participants). Comparison between Low/High PRS groups were carried out using Student t-test for continuous variables and chi-square test for categorical variables.

| Sample descriptive | Males |  |  |  | Females |  |  |  |
| --- | --- | --- | --- | --- | --- | --- | --- | --- |
|  | Total<br>(n =<br>249) | Low<br>PRS<br>(n =<br>124) | High<br>PRS<br>(n =<br>125) | *p* | Total<br>(n =<br>217) | Low<br>PRS<br>(n =<br>108) | High<br>PRS<br>(n =<br>109) | *p* |
| Age at delivery<br>(years) | 31.16<br>(4.86) | 31.21<br>(4.7) | 31.11<br>(5.03) | 0.880 | 31.1<br>(5.31) | 30.81<br>(4.8) | 31.38<br>(5.77) | 0.434 |
| Birth weight<br>(grams) | 3149<br>(427) | 3126<br>(439) | 3172<br>(415) | 0.398 | 3081<br>(427) | 3065<br>(421) | 3098<br>(433) | 0.570 |
| Breastfeeding at 3<br>months- yes | 61.6%<br>(146) | 60.2%<br>(71) | 63%<br>(75) | 0.750 | 55.2%<br>(117) | 59.8%<br>(64) | 50.5%<br>(53) | 0.219 |
| Smoking during<br>pregnancy | 2% (5) | 3.2% (4) | 0.8% (1) | 0.361 | 2.8% (6) | 2.8% (3) | 2.8% (3) | 1 |
| Household income <<br>SG\$2000 | 0.13<br>(0.34) | 0.1<br>(0.31) | 0.16<br>(0.37) | 0.201 | 0.16<br>(0.36) | 0.15<br>(0.36) | 0.17<br>(0.37) | 0.732 |
| Ethnicity |  |  |  | 0.944 |  |  |  | **0.016** |
| Chinese | 53.8%<br>(134) | 54%<br>(67) | 53.6%<br>(67) |  | 58.5%<br>(127) | 60.2%<br>(65) | 56.9%<br>(62) |  |
| Indian |  |  |  |  |  |  |  |  |
| Malay | 15.3% | 14.5% | 16% |  | 13.8% | 7.4% (8) | 20.2% |  |
|  | (38) | (18) | (20) |  | (30) | 32.4% | (22) |  |
|  | 30.9% | 31.5% | 30.4% |  | 27.6% | (35) | 22.9% |  |
|  | (77) | (39) | (38) |  | (60) |  | (25) |  |
| Gestational age at birth (weeks) | 38.77<br>(1.28) | 38.7<br>(1.3) | 38.84<br>(1.25) | 0.400 | 38.96<br>(1.21) | 38.89<br>(1.19) | 39.03<br>(1.22) | 0.395 |

**Table 3.**
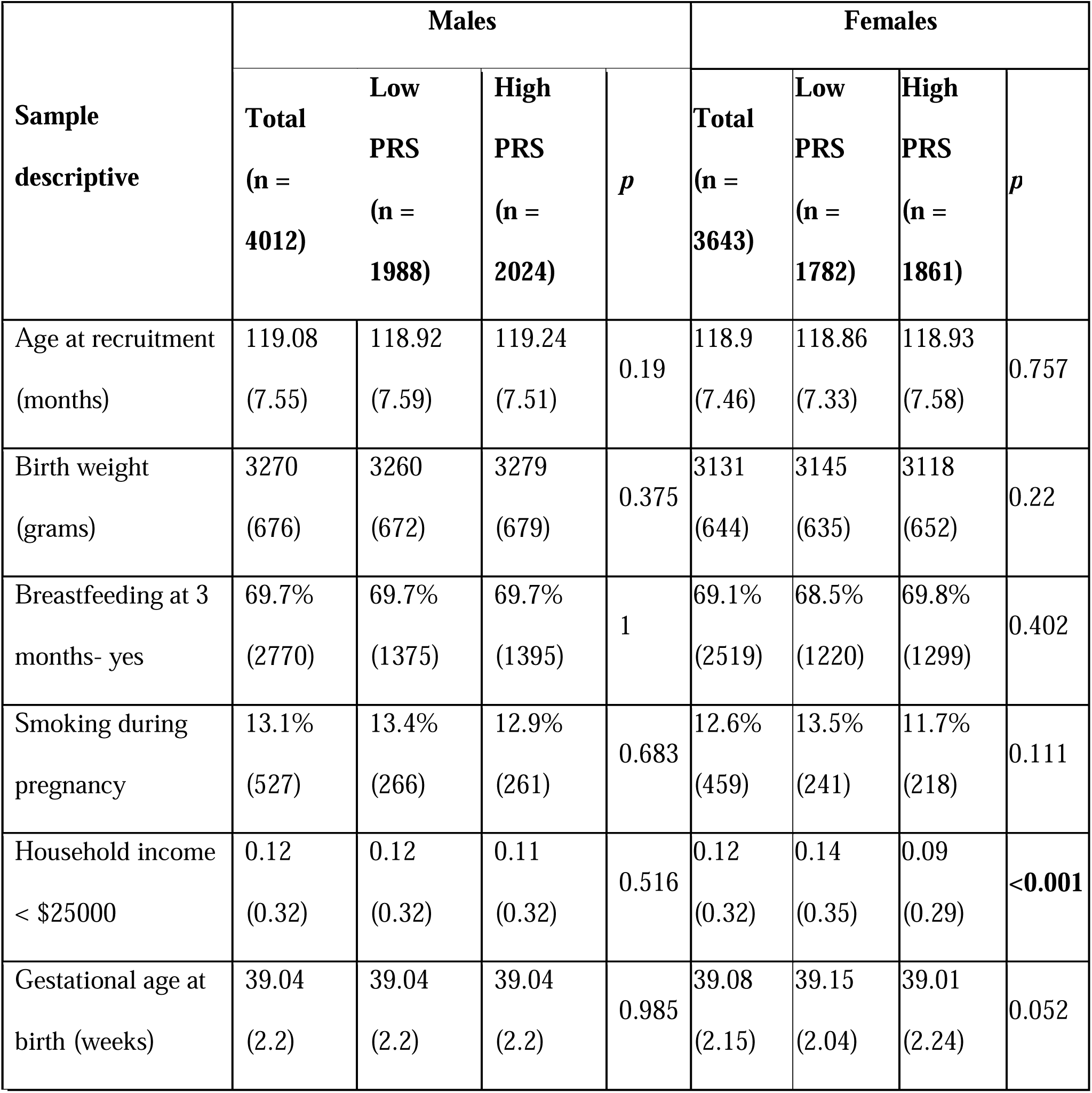
Participants’ characteristics in ABCD. Numbers are presented as mean (SD) or percentage (number of participants). Comparison between Low/High PRS groups were carried out using Student t-test for continuous variables and chi-square test for categorical variables.

**Table 4.**
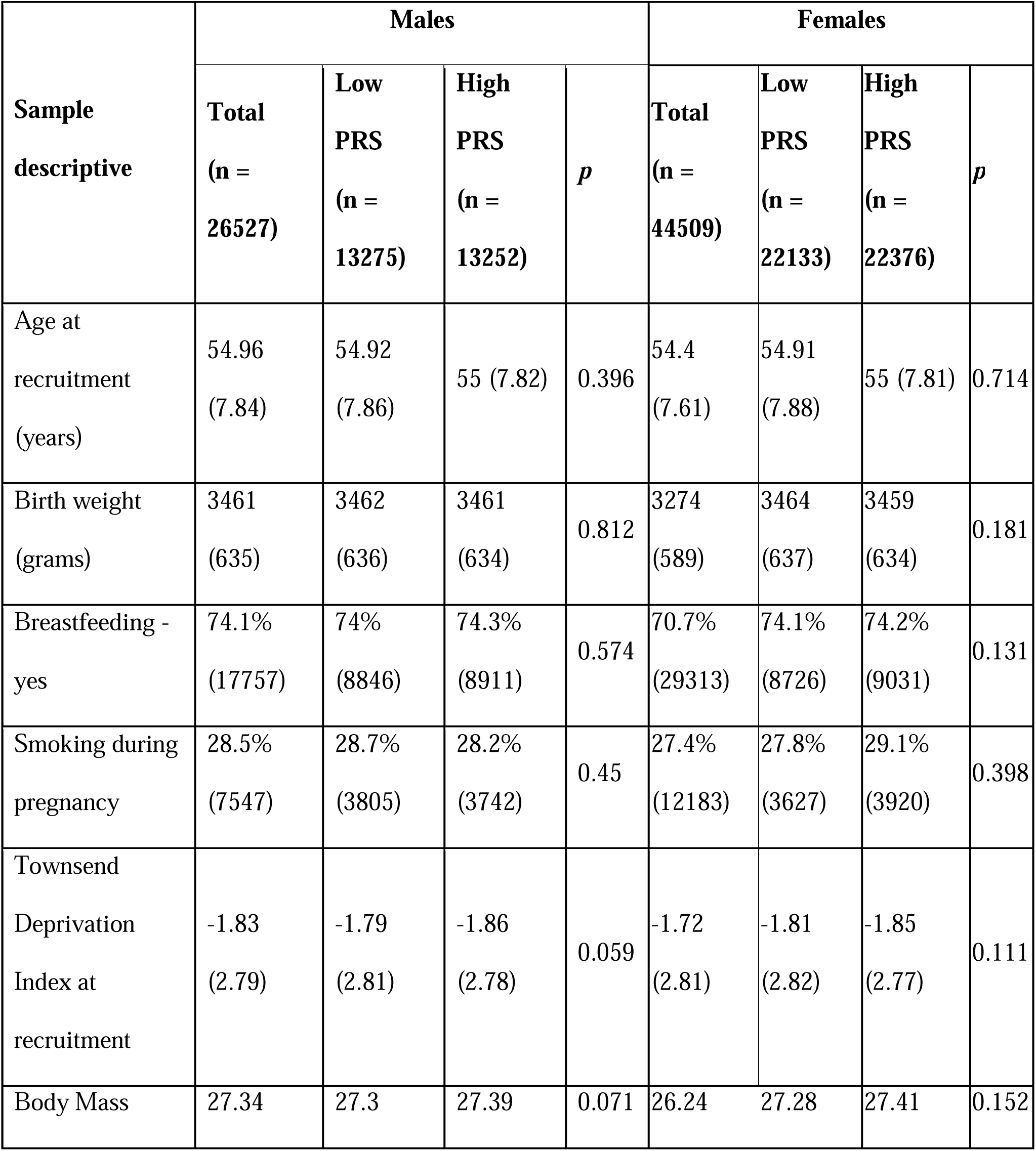
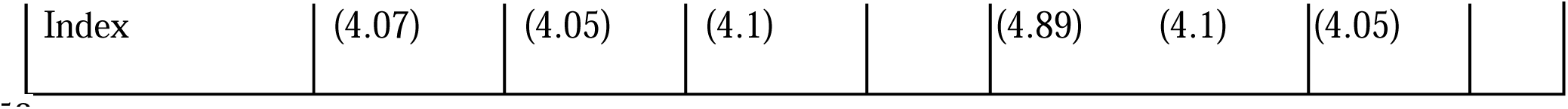
Participants’ characteristics in UKB. Numbers are presented as mean (SD) or percentage (number of participants). Comparison between Low/High PRS groups were carried out using Student t-test for continuous variables and chi-square test for categorical variables.

#### Enrichment Analysis

Enrichment analyses for gene ontologies were performed using MetaCore^TM^ (Clarivate Analytics) on the SNPs that compose the FI rPRS for males and females separately. Furthermore, gene-based enrichment analyses were performed in FUMA (https://fuma.ctglab.nl/) (Aguet *et al*, 2019; MacArthur *et al*, 2017; Watanabe *et al*, 2017) after mapping the SNPs composing the FI rPRS to genes with the biomaRt package in R (Durinck *et al*, 2005; Durinck *et al*, 2009).

### Investigating the causal relationship between high fasting insulin and altered executive functions according to early adversity exposure in a sex-specific manner

#### Two-Sample Mendelian Randomization Analyses

Two-sample MR analyses were performed using R (Durinck *et al*, 2005; Durinck *et al*, 2009) and the TwoSampleMR package (Hemani *et al*, 2018b). Exposure and outcome GWAS summary statistics were harmonized as described within the package. To assess horizontal pleiotropy (i.e., an association of the genetic instrument with the outcome independent of the exposure), we used the MR-Egger estimation for genetic instruments. When the pleiotropy assumption was not met, we used a different statistical method robust to pleiotropy: MR pleiotropy residual sum and outlier (MR-PRESSO). MR-PRESSO is able to identify outliers with potential horizontal pleiotropy when using multiple genetic variants as an instrument and provides a corrected estimate after removing these outliers (Zhao and Schooling, 2021). We first performed fixed-effects meta-analysis of genetic instruments using inverse-variance weighting (IVW) (Hemani *et al*, 2018a). Standard errors were computed with the Wald estimator and delta weighting to account for uncertainty in genetic association with the exposure. Then, to assess the robustness of our findings, we performed a simple median MR and weighted median MR. Since we were interested in inspecting the interaction effect between FI and adversity exposure on impulsivity, we separate analysis on males and females: 1) two-sample MR between the FI GWAS and an impulsivity GWAS constructed with data from individuals exposed to adversity; 2) two-sample MR between the FI GWAS and an impulsivity GWAS constructed with data from individuals that were not exposed to adversity.

## Results

### Investigating the shared genetic background between higher fasting insulin and altered executive function phenotypes (impulsivity and ADHD)

The conditional Q-Q plots did not show enrichment for FI given impulsivity or ADHD as evident in *supplementary Figure S8*. The blue lines in *supplementary Figure S8* are drawn using the FI GWAS including all the SNPs regardless of their association with impulsivity or ADHD. An increasingly leftward deflection from the dashed line of no association was not observed when SNPs with stronger association with impulsivity and ADHD were plotted. To provide a list of shared loci between FI and impulsivity or ADHD, we performed conjFDR analyses where we found no SNPs in common between the genetic datasets.

Summary statistics for the sex-specific impulsivity GWAS generated in this study can be found at https://github.com/SilveiraLab/Sex-Specific_Impulsivity_GWAS. A brief description of the findings from this GWAS can be found in the Supplementals (*Figures S1, S2, S3, S4, S5, and S6*).

### Investigating the interactive effects between polygenic risk scores for higher fasting insulin and childhood adversity on altered executive function phenotypes in a sex-specific manner

Baseline comparisons between low and high PRS groups were performed in all four cohorts. In the MAVAN cohort, the only main confounding variable that was significantly different between low and high PRS was birth weight in males (p = 0.034) (*Table 1).* In the GUSTO cohort, ethnicity in females (p = 0.016) was significantly different between the low and high PRS as shown in *Table 2*. In the ABCD cohort, household income in females (p < 0.001), was significantly different between low and high PRS (*Table 3)*. In the UKB cohort, there were no significantly different confounding variables between low and high PRS (*Table 4)*.

#### PheWAS Analysis

We performed linear and logistic regression analyses to investigate the interaction between rPRS for FI and postnatal adversity exposure on EF behavior separately in males and females across MAVAN, GUSTO, ABCD, and UKB cohort. Analyses were adjusted by population stratification PCs, age, and other covariates if they were found to be significantly different between low/high PRS groups (*Tables 1–4*). Significant interaction effects, presented by nominal p-values in each cohort, are highlighted in *Figure 1*. None of the outcomes remained significantly were associated after adjusting for multiple testing. Results for the enrichment analyses can be found in supplementary materials.

**Figure 1.**
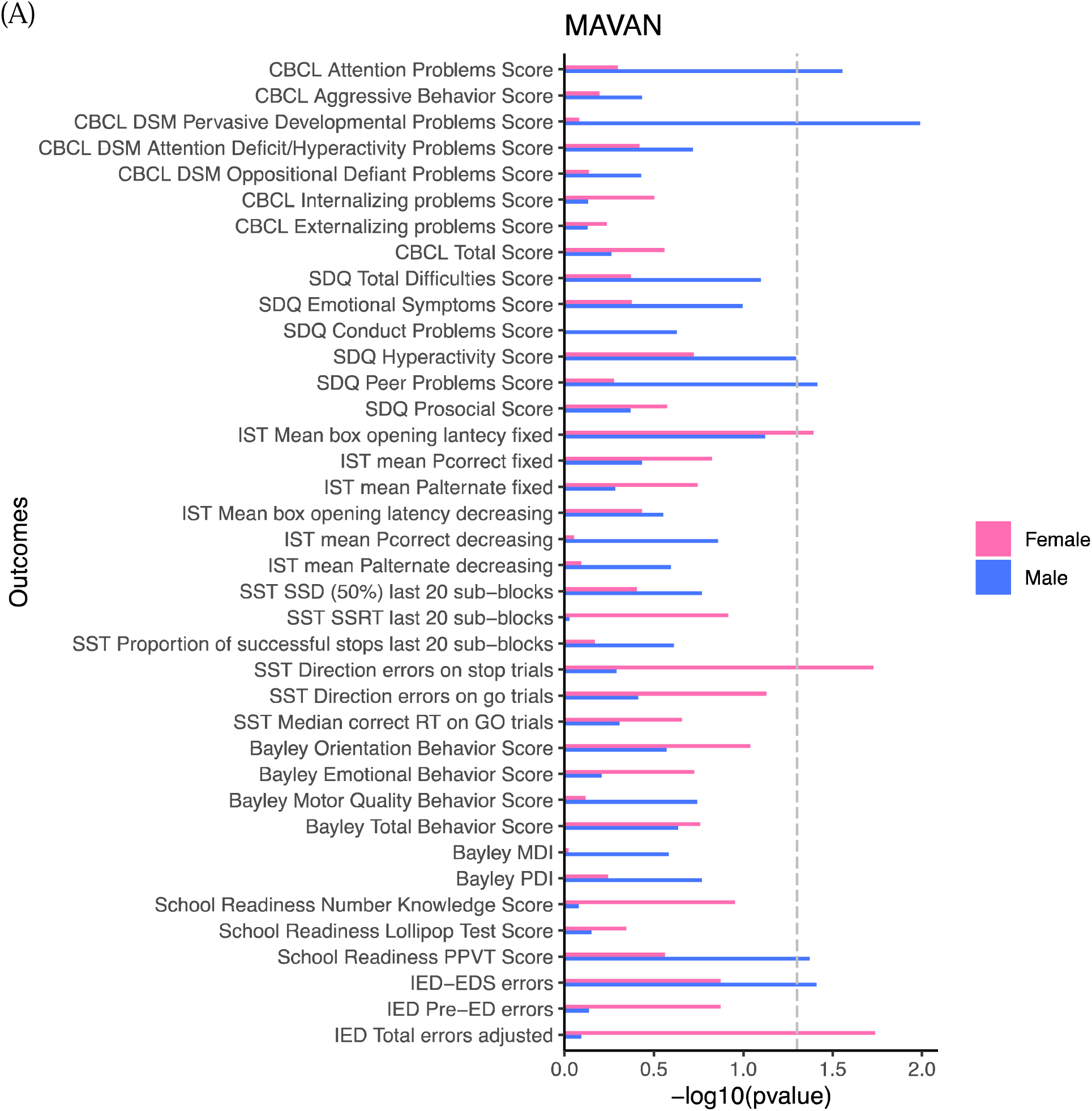
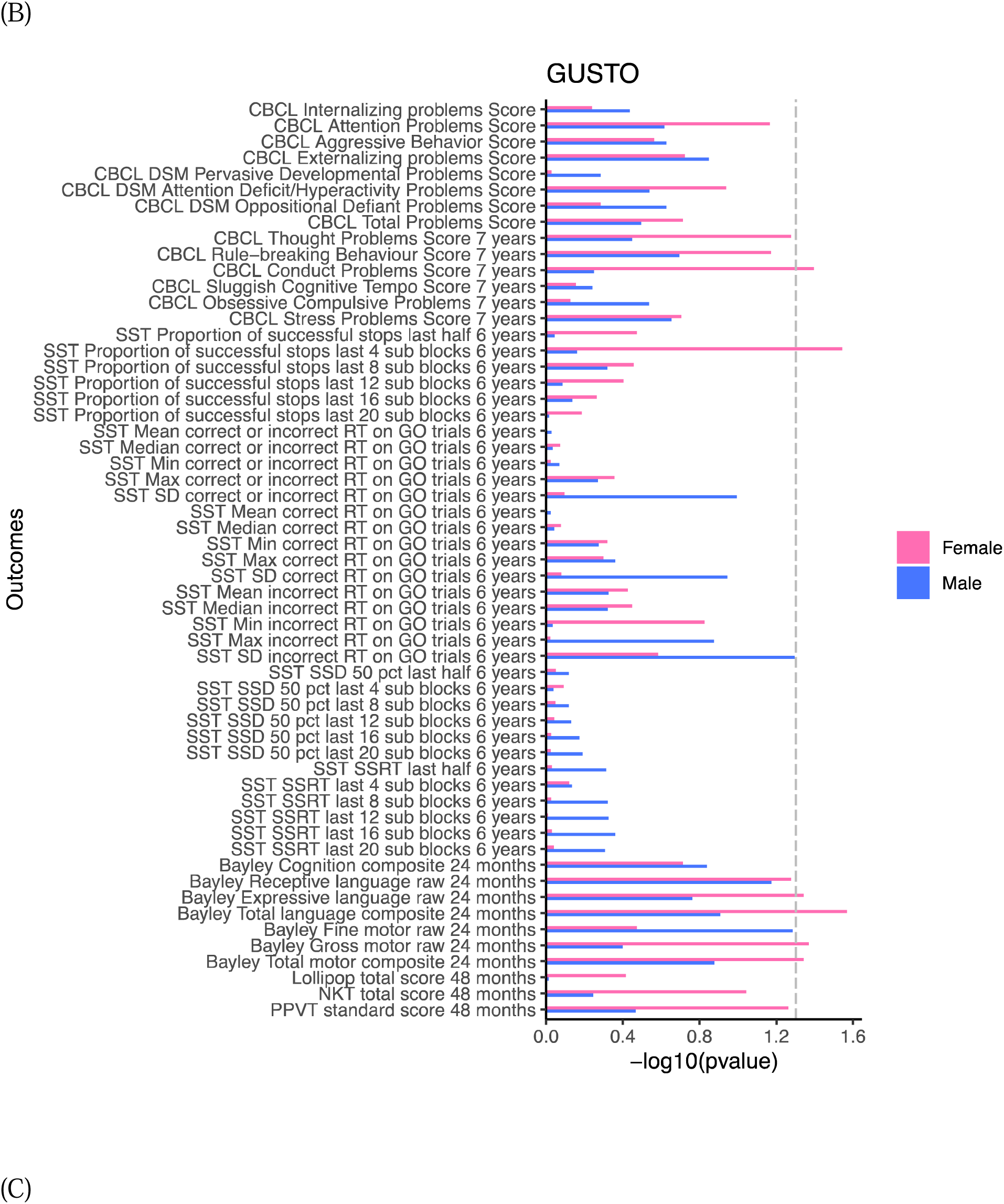
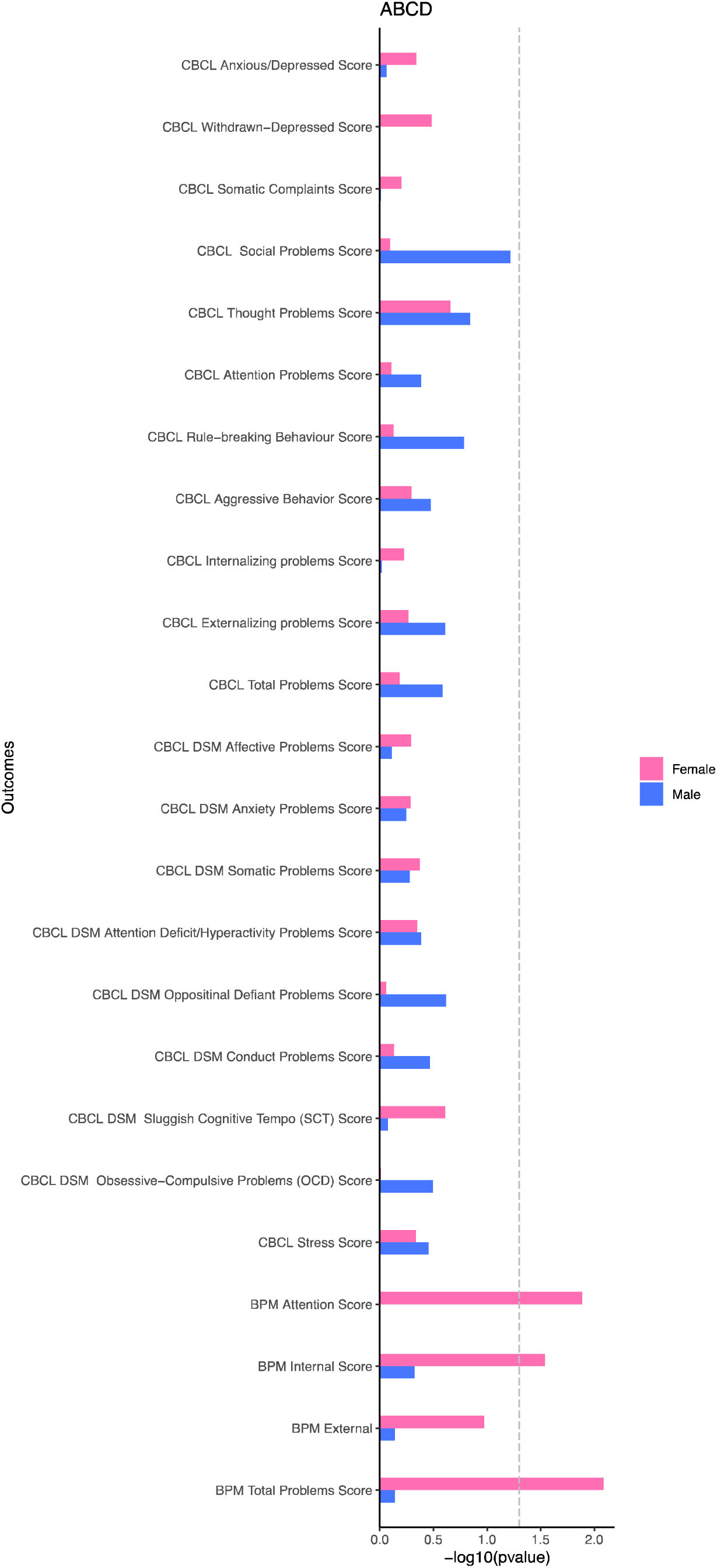
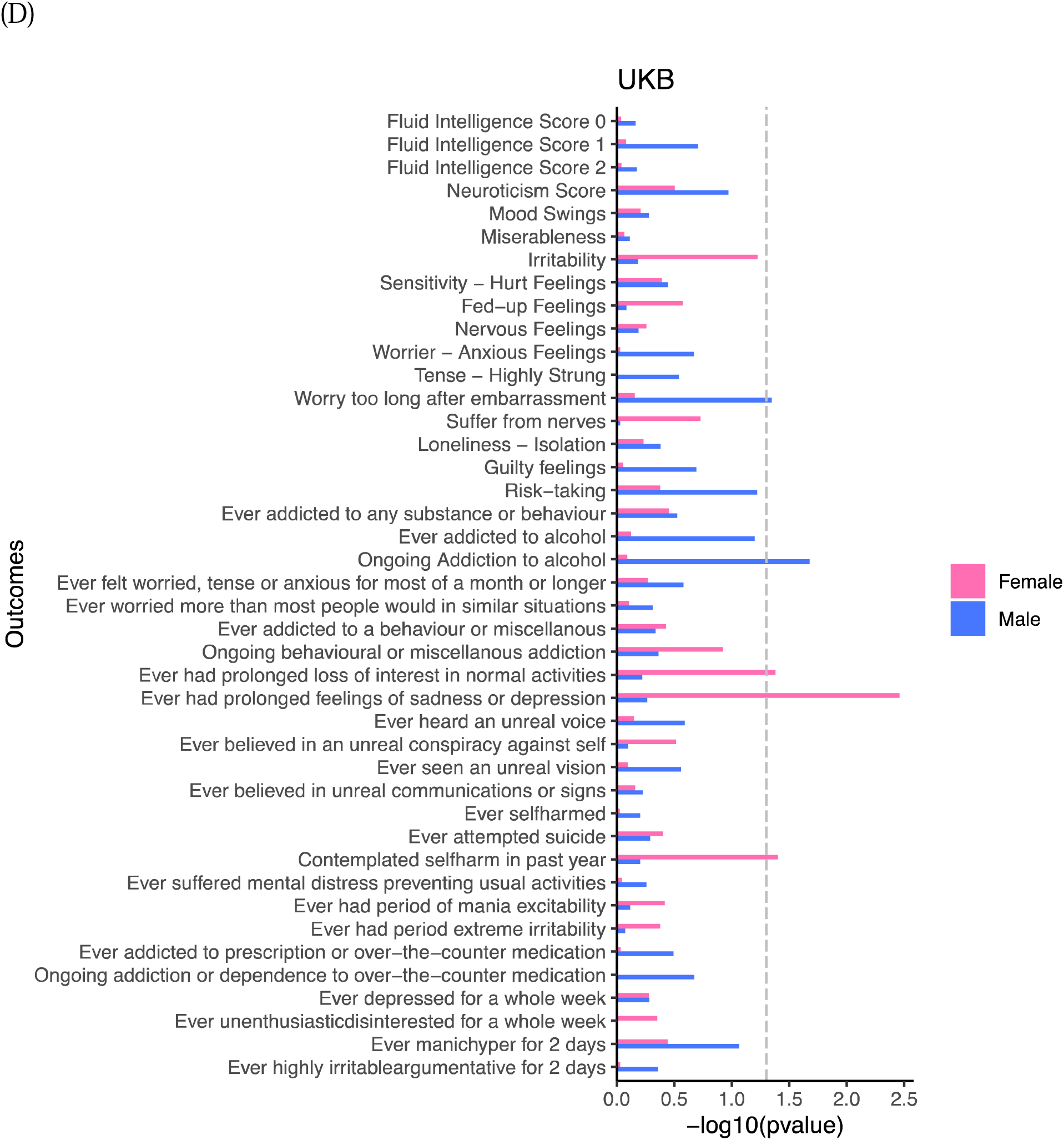
Executive functions PheWAS Results. Linear/logistic regression analyses were done to investigate the interaction effect between the rPRS for FI and postnatal adversity exposure on executive function behaviors in MAVAN (A), GUSTO (B), ABCD (C), and UKB (D), adjusted by covariates described in the methods section. In all cohorts, there were no significant interaction effects found on executive functions when considering the adjusted FDR p-values. The results highlight outcomes that were significant before the FDR correction was applied.

### Investigating the interactive effects between polygenic scores for higher fasting insulin and childhood adversity on altered executive function phenotypes in a sex-specific manner

Mendelian randomization analyses tested potential causal associations between FI and impulsivity. Before running two-sample MR, we used the MR Egger intercept test to verify horizontal pleiotropy. Test results did not reject the hypothesis of horizontal pleiotropy in males exposed to adversity (intercept = –0.00008; standard error = 0.00002; p < 0.001), in males not exposed to adversity (intercept = 0.00009; standard error = 0.00001; p < 0.001), and in females not exposed to adversity (intercept = 0.00004; standard error = 0.00001; p = 0.001). Using MR-PRESSO, we identified and remove outliers in those three cases and performed MR Egger again. The test could not significantly deny the presence of horizontal pleiotropy in males exposed to adversity (intercept = –0.0001; standard error = 0.00002; p < 0.001), in males not exposed to adversity (intercept = 0.00008; standard error = 0.00001; p < 0.001), and in females not exposed to adversity (intercept = 0.00003; standard error = 0.00001; p = 0.017).

Interestingly, the MR Egger intercept test suggested no horizontal pleiotropy in females exposed to adversity (intercept = –0.000006; standard error = 0.00002; p = 0.766). Therefore, two-sample MR analysis was used to inspect the potential causal association between FI and impulsivity in females exposed to adversity. All MR methods resulted in a significant association between the exposure FI and the outcome impulsivity in females exposed to adversity [IVW p < 0.001, IVW β = –0.028, IVW 95% CI: –0.032 to –0.023; Simple Median p < 0.001, Simple Median β = –0.035, Simple Median 95% CI: –0.042 to –0.029; Weighted Median p < 0.001, Weighted Median β = –0.022, Weighted Median 95% CI: –0.029 to –0.015].

## Discussion

This study aimed to investigate if FI had a moderating effect on the association between ELA and altered EF in a sex-specific manner, and whether there was a causal relationship between FI and altered EF according to adversity exposure.

Previous work from our lab showed that a mesocorticolimbic-specific expression based PRS of insulin receptor (IR-ePRS) was associated with childhood impulsivity in males (Dass *et al*, 2019). Based on these findings, we expected that insulin and impulsivity would share genetic architecture. However, our study could not confirm this neither in males nor females. CondFDR findings did not indicate a shared genetic architecture between FI and impulsivity, an EF previously associated with the interaction between high FI levels and ELA in (Batra *et al*, 2021), nor between FI and ADHD, a disorder in which impulsivity is a symptom (Kenemans *et al*, 2005). The lack of shared genetic architecture suggests there may be factor interacting with FI (e.g. ELA) resulting in altered EF behaviors. These findings led us to investigate the interaction between FI and ELA on EF behaviors, including a broad range of outcomes.

We demonstrated that the calculation of a rPRS, consisting of SNPs most highly associated with peripheral insulin levels in children and representing the risk for high FI levels early in life, can be derived from the GWAS of FI in adults (Batra *et al*, 2021). Insulin resistance has been shown to be associated with childhood social relationship problems (Gangel *et al*, 2020), and these may also affect EF development, as observed in our results. Since conduct problems often occur with ADHD (Faraone, 2000) and heritability has been established in molecular genetic studies between both traits (Holmes *et al*, 2002), it made sense that we observed a significant interaction effects on conduct problems and rule breaking behavior in GUSTO females. While in MAVAN, GUSTO, and ABCD, attention and impulsivity traits were being highlighted through this interaction effect using different measures, in the UKB, the significant interaction effect was seen on psychiatric outcomes, such as addiction to alcohol in males and depression in females. The significant outcomes in the UKB show a trend from childhood to adulthood because impulsivity and attention related problems map to addiction (Kreek *et al*, 2005) and depression (Granö *et al*, 2007) in adulthood. Additionally, internalizing problems are significantly associated with symptoms of anxiety and depression (Pedersen *et al*, 2021), confirming results that we saw as early as adolescence in ABCD females.

Although none of these interaction effects were significant after adjusting for multiple testing, the trend is present throughout development in both males and females during childhood, as evident through MAVAN and GUSTO. However, is adolescence, this association becomes only significant in females in the ABCD cohort. This may be explained by males exhibiting higher insulin sensitivity at this age, serving as a metabolic corrector (Hoffman *et al*, 2000). When examined in adults, both analyses in males and females showed significant interaction between FI and ELA on psychiatric outcomes, thereby reaffirming the trend of insulin functioning’s association with cognitive functions (Ma *et al*, 2015) in childhood, which has been shown in literature and in our studies.

The gene ontology enrichment analyses, done through MetaCore^TM^, showed that the SNPs composing the FI rPRS are associated with nervous system development. Some of the enriched processes should be highlighted, such as dopamine D2 receptor transactivation in males as the dopamine system has been linked to impulsive behavior in animal models and human studies (Dalley and Roiser, 2012; van Gaalen *et al*, 2006). This finding suggests a potential neurodevelopment pathway to support the relationship between the genetic background linked to insulin, ELA, and dopamine. For example, in previous animal studies from our lab, rats who have experienced ELA display displayed dopamine release in response to palatable food, an effect which is reverted by insulin administration (Laureano *et al*, 2019). These findings highlight that the relationship between ELA and dopamine is moderated by insulin. Further, enrichment analyses in FUMA revealed that the genes, to which the SNPs composing the FI rPRS map to, were significantly enriched for cognition and ADHD in both males and females for several GWASs, and for impulsivity in females. These findings align with our executive function results from the interaction analysis.

MR is a powerful tool as it uses Mendel’s law to assess the potential causal association between two traits through a naturally occurring randomized clinical trial (Kappelmann *et al*, 2021). In our study, MR showed significant associations between FI and impulsivity in females exposed to adversity. In all other cases, the Egger test and the MR-PRESSO test failed to significantly deny the presence of horizontal pleiotropy. In other words, there may be causal association between FI and impulsivity in those cases, but MR could not be used to determine its presence. These results are consistent with previous studies from our lab where ELA has specifically affected female behavior. Barbieri et al (Barbieri *et al*, 2009a) showed that women born with severe intrauterine growth (IUGR) displayed a preference for carbohydrates in adulthood compared to those born with normal birth weight, without effects in men (Barbieri *et al*, 2009a). IUGR has been associated with metabolic stress which can result in altered appetite regulation. Appetite regulation involves the EF inhibition of impulsive behaviors (Gluck *et al*, 2017) explaining the results from Barbieri et al in the context of our study. A similar effect was observed in another study performed by our lab, whereby IUGR girls demonstrated increased impulsivity in the Snack delay task, while there were no effects of IUGR in boys (Silveira *et al*, 2012). Rodent studies done on the topic also demonstrate that ELA leads to learning impairments in female rats, and prepubertal stress leads to compulsive behavior in female rats, an effect that is not observed in males. (Brydges *et al*, 2015). Overall, we can conclude that there is a causal association between insulin and impulsivity in the context of adversity in females. Additional investigations are needed to exclude the possibility of this causal relationship existing in males.

## Conclusion

In conclusion, our current findings offer evidence for how FI moderates the long-term effects of ELA on altered EF, specifically impulsivity and attention, in males and females throughout development. High FI also appears to be causally associated with increased impulsivity in females exposed to adversity. This study is impactful in highlighting the mechanisms by which insulin moderated the effects of ELA on EF disorders, and it provides insights into potential mitigating interventions.

## Data Availability

All data in the present work are contained in the Supplementary Materials.

## Acknowledgements

We are extremely grateful to the families who took part in this study, the midwives for their help in recruiting them, and the entire ALSPAC team, including interviewers, computer and laboratory technicians, clerical workers, research scientists, volunteers, managers, receptionists and nurses.

This research was supported by: Canadian Institutes of Health Research (CIHR, PJT-166066, PPS and IP), the JPB Foundation through a grant to the JPB Research Network on Toxic Stress: A Project of the Center on the Developing Child at Harvard University, Fonds de recherche du Québec – Santé (FRQS to AB and PPS), awarded for the project: Les variations de la function de l’insuline modulent l’adversité au début de la vie sur les compartements liés à la dopamine mésocorticolimbique. Finally, this research was also supported by an award from the McGill-Douglas Max Planck Institute of Psychiatry International Collaborative Initiative in Adversity and Mental Health, an international partnership funded by the Healthy Brains for Healthy Lives initiative. The UK Medical Research Council and Wellcome (Grant ref: 217065/Z/19/Z) and the University of Bristol provide core support for ALSPAC. This publication is the work of the authors and will serve as guarantors for the contents of this paper. A comprehensive list of grants funding is available on the ALSPAC website (http://www.bristol.ac.uk/alspac/external/documents/grant-acknowledgements.pdf); ALSPAC GWAS data was generated by Sample Logistics and Genotyping Facilities at Wellcome Sanger Institute and LabCorp (Laboratory Corporation of America) using support from 23andMe.

The authors declare no competing interest.

## Notes

### Competing Interest Statement

The authors have declared no competing interest.

### Author Declarations

Ethics approval for the study was obtained from the ALSPAC Ethics and Law Committee and the local research ethics committees. Approval for the MAVAN project was obtained by the ethics committees and university affiliates (McGill University and Universite de Montreal, the Royal Victoria Hospital, Jewish General Hospital, Centre hospitalier de lUniversite de Montreal and Hopital Maisonneuve-Rosemount) and St. Josephs Hospital and McMaster University, Hamilton, Ontario, Canada. This study was approved by the National University Hospital and KK Womens and Childrens Hospital, National Healthcare Group Domain Specific Review Board (NHG DSRB Ref D/09/021) and Sing Health Centralized Institutional Review Board (CIRB Ref 2018/2767). The institutional review board of the University of California, San Diego centralized the approval for data collection from ABCD. Approval for the UK Biobank was obtained by the North West Multicentre Research 580 Ethics Committee, the National Information Governance Board for Health and Social Care and the Community Health Index Advisory Group.

## References

1. Achenbach TM, Edelbrock C (1991). Child behavior checklist. Burlington (Vt) 7: 371–392.

2. Achenbach TM, McConaughy S, Ivanova M, Rescorla L (2011). Manual for the ASEBA brief problem monitor (BPM). Burlington, VT: ASEBA 33.

3. Aguet F, Barbeira AN, Bonazzola R, Jo B, Kasela S, Liang Y, et al (2019). The GTEx Consortium Atlas of Genetic Regulatory Effects Across Human Tissues. Science (New York, NY) 369(6509): 1318–1330.

4. Andreassen OA, Djurovic S, Thompson WK, Schork AJ, Kendler KS, O’Donovan MC, et al (2013a). Improved detection of common variants associated with schizophrenia by leveraging pleiotropy with cardiovascular-disease risk factors. The American Journal of Human Genetics 92(2): 197–209.

5. Andreassen OA, Thompson WK, Dale AM (2014). Boosting the power of schizophrenia genetics by leveraging new statistical tools. Schizophrenia bulletin 40(1): 13–17.

6. Andreassen OA, Thompson WK, Schork AJ, Ripke S, Mattingsdal M, Kelsoe JR, et al (2013b). Improved detection of common variants associated with schizophrenia and bipolar disorder using pleiotropy-informed conditional false discovery rate. PLoS genetics 9(4): e1003455.

7. Balasundaram P, Avulakunta ID (2021). Bayley scales of infant and toddler development. StatPearls [Internet]. StatPearls Publishing.

8. Barbieri MA, Portella AK, Silveira PP, Bettiol H, Agranonik M, Silva AA, et al (2009a). Severe intrauterine growth restriction is associated with higher spontaneous carbohydrate intake in young women. Pediatric research 65(2): 215–220.

9. Barbieri MA, Portella AK, Silveira PP, Bettiol H, Agranonik M, Silva AA, et al (2009b). Severe intrauterine growth restriction is associated with higher spontaneous carbohydrate intake in young women. Pediatric research 65(2): 215–220.

10. Batra A, Chen LM, Wang Z, Parent C, Pokhvisneva I, Patel S, et al (2021). Early Life Adversity and Polygenic Risk for High Fasting Insulin Are Associated With Childhood Impulsivity. Frontiers in neuroscience 15: 704785.

11. Bøe T, Hysing M, Skogen JC, Breivik K (2016). The Strengths and Difficulties Questionnaire (SDQ): Factor structure and gender equivalence in Norwegian adolescents. PloS one 11(5): e0152202.

12. Boyd A, Golding J, Macleod J, Lawlor DA, Fraser A, Henderson J, et al (2013). Cohort Profile: The ‘Children of the 90s’—The Index Offspring of the Avon Longitudinal Study of Parents and Children. Int J Epidemiol 42(1): 111–127.

13. Brydges NM, Holmes MC, Harris AP, Cardinal RN, Hall J (2015). Early life stress produces compulsive-like, but not impulsive, behavior in females. Behavioral neuroscience 129(3): 300.

14. Chen Q, Hartman CA, Haavik J, Harro J, Klungsøyr K, Hegvik T-A, et al (2018). Common psychiatric and metabolic comorbidity of adult attention-deficit/hyperactivity disorder: A population-based cross-sectional study. PloS one 13(9): e0204516.

15. Chikazoe J, Jimura K, Hirose S, Yamashita K-i, Miyashita Y, Konishi S (2009). Preparation to inhibit a response complements response inhibition during performance of a stop-signal task. Journal of Neuroscience 29(50): 15870–15877.

16. Cyders MA (2013). Impulsivity and the sexes: Measurement and structural invariance of the UPPS-P Impulsive Behavior Scale. Assessment 20(1): 86–97.

17. Dalley JW, Roiser J (2012). Dopamine, Serotonin and Impulsivity. Neuroscience 215: 42–58.

18. Dass SAH, McCracken K, Pokhvisneva I, Chen LM, Garg E, Nguyen TT, et al (2019). A Biologically-Informed Polygenic Score Identifies Endophenotypes and Clinical Conditions Associated with the Insulin Receptor Function on Specific Brain Regions. E Bio Medicine 42: 188–202.

19. Durinck S, Moreau Y, Kasprzyk A, Davis S, De Moor B, Brazma A, et al (2005). BioMart and Bioconductor: A Powerful Link Between Biological Databases and Microarray Data Analysis. Bioinformatics 21(16): 3439–3440.

20. Durinck S, Spellman PT, Birney E, Huber W (2009). Mapping Identifiers for the Integration of Genomic Datasets with the R/Bioconductor Package biomaRt. Nat Protoc 4(8): 1184.

21. Faraone SV (2000). Genetics of childhood disorders: XX. ADHD, Part 4: is ADHD genetically heterogeneous? Journal of the American Academy of Child & Adolescent Psychiatry.

22. Fraser A, Macdonald-Wallis C, Tilling K, Boyd A, Golding J, Davey Smith G, et al (2013). Cohort Profile: the Avon Longitudinal Study of Parents and Children: ALSPAC Mothers Cohort. Int J Epidemiol 42(1): 97–110.

23. Gan Y, Meng L, Xie J (2016). Comparison of school readiness between rural and urban Chinese preschool children. Social Behavior and Personality: an international journal 44(9): 1429–1442.

24. Gangel MJ, Dollar J, Brown A, Keane S, Calkins SD, Shanahan L, et al (2020). Childhood social preference and adolescent insulin resistance: Accounting for the indirect effects of obesity. Psychoneuroendocrinology 113: 104557.

25. Ghasemi R, Haeri A, Dargahi L, Mohamed Z, Ahmadiani A (2013). Insulin in the Brain: Sources, Localization and Functions. Mol Neurobiol 47(1): 145–171.

26. Gluck ME, Viswanath P, Stinson EJ (2017). Obesity, appetite, and the prefrontal cortex. Current obesity reports 6(4): 380–388.

27. Granö N, Keltikangas-Järvinen L, Kouvonen A, Virtanen M, Elovainio M, Vahtera J, et al (2007). Impulsivity as a predictor of newly diagnosed depression. Scandinavian journal of psychology 48(2): 173–179.

28. Hemani G, Bowden J, Davey Smith G (2018a). Evaluating the potential role of pleiotropy in Mendelian randomization studies. Human molecular genetics 27(R2): R195–R208.

29. Hemani G, Zheng J, Elsworth B, Wade KH, Haberland V, Baird D, et al (2018b). The MR-Base platform supports systematic causal inference across the human phenome. elife 7.

30. Heni M, Kullmann S, Preissl H, Fritsche A, Häring H-U (2015). Impaired Insulin Action in the Human Brain: Causes and Metabolic Consequences. Nat Rev Endocrinol 11(12): 701.

31. Hoffman RP, Vicini P, Sivitz WI, Cobelli C (2000). Pubertal adolescent male-female differences in insulin sensitivity and glucose effectiveness determined by the one compartment minimal model. Pediatric research 48(3): 384–388.

32. Holmes J, Payton A, Barrett J, Harrington R, McGuffin P, Owen M, et al (2002). Association of DRD4 in children with ADHD and comorbid conduct problems. American Journal of Medical Genetics 114(2): 150–153.

33. Honeycutt JA, Demaestri C, Peterzell S, Silveri MM, Cai X, Kulkarni P, et al (2020). Altered corticolimbic connectivity reveals sex-specific adolescent outcomes in a rat model of early life adversity. Elife 9.

34. Jernigan TL, Brown SA, Dowling GJ (2018). The adolescent brain cognitive development study. Journal of research on adolescence: the official journal of the Society for Research on Adolescence 28(1): 154.

35. Kappelmann N, Arloth J, Georgakis MK, Czamara D, Rost N, Ligthart S, et al (2021). Dissecting the association between inflammation, metabolic dysregulation, and specific depressive symptoms: a genetic correlation and 2-sample Mendelian randomization study. JAMA psychiatry 78(2): 161–170.

36. Kenemans J, Bekker E, Lijffijt M, Overtoom C, Jonkman L, Verbaten M (2005). Attention deficit and impulsivity: selecting, shifting, and stopping. International Journal of Psychophysiology 58(1): 59–70.

37. Kettunen J, Demirkan A, Würtz P, Draisma HH, Haller T, Rawal R, et al (2016). Genome-wide study for circulating metabolites identifies 62 loci and reveals novel systemic effects of LPA. Nature communications 7(1): 1–9.

38. Kreek MJ, Nielsen DA, Butelman ER, LaForge KS (2005). Genetic influences on impulsivity, risk taking, stress responsivity and vulnerability to drug abuse and addiction. Nature neuroscience 8(11): 1450–1457.

39. Kuijpers RC, Otten R, Krol NP, Vermulst AA, Engels RC (eds) (2013). The reliability and validity of the Dominic Interactive: A computerized child report instrument for mental health problems. Child & Youth Care Forum. Springer.

40. Kullmann S, Heni M, Hallschmid M, Fritsche A, Preissl H, Häring H-U (2016). Brain Insulin Resistance at the Crossroads of Metabolic and Cognitive Disorders in Humans. Physiol Rev 96(4): 1169–1209.

41. Lagou V, Mägi R, Hottenga J-J, Grallert H, Perry JR, Bouatia-Naji N, et al (2021). Sex-dimorphic genetic effects and novel loci for fasting glucose and insulin variability. Nature communications 12(1): 1–18.

42. Laureano D, Alves M, Miguel P, Machado T, Reis A, Mucellini A, et al (2019). Intrauterine Growth Restriction Modifies the Accumbal Dopaminergic Response to Palatable Food Intake. Neuroscience 400: 184–195.

43. Ma L, Wang J, Li Y (2015). Insulin resistance and cognitive dysfunction. Clinica chimica acta 444: 18–23.

44. MacArthur J, Bowler E, Cerezo M, Gil L, Hall P, Hastings E, et al (2017). The New NHGRI-EBI Catalog of Published Genome-Wide Association Studies (GWAS Catalog). Nucleic Acids Res 45(D1): D896–D901.

45. Northstone K, Lewcock M, Groom A, Boyd A, Macleod J, Timpson N, et al (2019). The Avon Longitudinal Study of Parents and Children (ALSPAC): An Update on the Enrolled Sample of Index Children in 2019. Wellcome Open Res 4.

46. O’Donnell KA, Gaudreau H, Colalillo S, Steiner M, Atkinson L, Moss E, et al (2014). The Maternal Adversity, Vulnerability and Neurodevelopment Project: Theory and Methodology. Can J Psychiatry 59(9): 497–508.

47. O’Connell KS, Shadrin A, Smeland OB, Bahrami S, Frei O, Bettella F, et al (2020). Identification of genetic loci shared between attention-deficit/hyperactivity disorder, intelligence, and educational attainment. Biological psychiatry 87(12): 1052–1062.

48. Pedersen ML, Jozefiak T, Sund AM, Holen S, Neumer S-P, Martinsen KD, et al (2021). Psychometric properties of the Brief Problem Monitor (BPM) in children with internalizing symptoms: examining baseline data from a national randomized controlled intervention study. BMC psychology 9(1): 1–12.

49. Pyykkönen A-J, Räikkönen K, Tuomi T, Eriksson JG, Groop L, Isomaa B (2010). Stressful life events and the metabolic syndrome: the prevalence, prediction and prevention of diabetes (PPP)-Botnia Study. Diabetes care 33(2): 378–384.

50. Sahakian BJ, Owen A (1992). Computerized assessment in neuropsychiatry using CANTAB: discussion paper. Journal of the Royal Society of medicine 85(7): 399.

51. Schuur M, Henneman P, Van Swieten J, Zillikens M, de Koning I, Janssens A, et al (2010). Insulin-resistance and metabolic syndrome are related to executive function in women in a large family-based study. European journal of epidemiology 25(8): 561–568.

52. Silveira PP, Agranonik M, Faras H, Portella AK, Meaney MJ, Levitan RD (2012). Preliminary Evidence for an Impulsivity-Based Thrifty Eating Phenotype. Pediatr Res 71(3): 293–298.

53. Silveira PP, Pokhvisneva I, Gaudreau H, Rifkin-Graboi A, Broekman BF, Steiner M, et al (2018). Birth weight and catch up growth are associated with childhood impulsivity in two independent cohorts. Scientific Reports 8(1): 1–10.

54. Silveira PP, Pokhvisneva I, Parent C, Cai S, Rema ASS, Broekman BF, et al (2017). Cumulative Prenatal Exposure to Adversity Reveals Associations with a Broad Range of Neurodevelopmental Outcomes that are Moderated by a Novel, Biologically Informed Polygenetic Score Based on the Serotonin Transporter Solute Carrier Family C6, Member 4 (SLC6A4) Gene Expression. Dev Psychopathol 29(5): 1601–1617.

55. Soh S-E, Tint MT, Gluckman PD, Godfrey KM, Rifkin-Graboi A, Chan YH, et al (2014). Cohort profile: Growing Up in Singapore Towards healthy Outcomes (GUSTO) birth cohort study. International journal of epidemiology 43(5): 1401–1409.

56. Sudlow C, Gallacher J, Allen N, Beral V, Burton P, Danesh J, et al (2015). UK biobank: an open access resource for identifying the causes of a wide range of complex diseases of middle and old age. PLoS medicine 12(3): e1001779.

57. van Gaalen MM, van Koten R, Schoffelmeer AN, Vanderschuren LJ (2006). Critical Involvement of Dopaminergic Neurotransmission in Impulsive Decision Making. Biol Psychiatry 60(1): 66–73.

58. Watanabe K, Taskesen E, Van Bochoven A, Posthuma D (2017). Functional Mapping and Annotation of Genetic Associations with FUMA. Nat Commun 8(1).

59. Zhao JV, Schooling CM (2021). Sex-specific associations of sex hormone binding globulin with CKD and kidney function: a univariable and multivariable Mendelian randomization study in the UK Biobank. Journal of the American Society of Nephrology 32(3): 686–694.

60. Zhao W-Q, Alkon DL (2001). Role of Insulin and Insulin Receptor in Learning and Memory. Mol Cell Endocrinol 177(1-2): 125–134.

