## Supplemental Materials for "Insulin moderates the effects of early life adversity on executive functioning in a sex-specific manner"

### Methods and Materials

#### Investigating the shared genetic background between higher fasting insulin and altered executive function phenotypes (impulsivity and ADHD)

*GWASs*

*Sex specific FI GWAS:* We obtained complete FI GWAS results in the form of summary statistics p-value from the Meta-Analyses of Glucose and Insulin-related traits Consortium (1). These GWAS meta-analysis results are provided for 47,806 men and 50,404 women. All participants were of European ancestry, without diabetes and mostly adults while data from a total of 8,222 adolescents were included in the meta-analyses. The additive genetic effect of each SNP was estimated using a linear regression model adjusting for age, study site, and genetic principal components. More information on this dataset can be found in Lagou et al (1).

*Sex specific Impulsivity GWAS:* As this GWAS was not available in the literature, we have performed a sex specific GWAS for impulsivity using the UK Biobank data. Genotyping data in the UKB cohort was available for 487,409 subjects. We excluded participants who withdrew their consent, with inconsistencies in genetic and reported sex, as well as outliers for heterozygosity. Also, we restricted our analysis only to the participants who identified themselves as "Caucasians" (ID 22006). We also removed variants with minor allele frequency < 0.01, an imputation accuracy info score < 0.1 as well as duplicated and ambiguous SNPs, resulting in 7,351,435 variants in the data set. The impulsivity phenotype was defined using mean time to correctly identify matches (ID 20023), obtained at initial assessment visit (2006-2010) during a reaction time test based on the card-game “Snap”. This variable measures the combined processing and reaction speed of a participant. There were 128,277 male and 146,607 female unrelated subjects with the impulsivity phenotype and genotype data available and included in the GWAS, as displayed in *Figures S1 and S2* respectively. Additionally, *Figures S3 and S4* show Q-Q plots of the observed P-values on those expected using all the variants analyzed in UK Biobank for males and females respectively. We applied linear regression analysis using SNPTEST v2.5.4 to assess the effect of each SNP on impulsivity, adjusting for age, genotyping array, and 40 genetic principal components.

As this is an original GWAS, a small description of the findings is provided. Gene- and region-based analyses of the significant genes ($P < 2.6*10^{-6}$) were conducted using MAGMA (Multi-marker Analysis of GenoMic Annotation) available on FUMA GWAS (Functional Mapping and Annotation of Genome-Wide Association Studies) (2). For gene-set pathway analysis we used the results obtained from the gene-based analysis considering SNPs at $10^{-5}$ as the threshold to conduct a further gene-set pathway analysis to test for gene enrichment using FUMA GWAS (Functional Mapping and Annotation of Genome-Wide Association Studies), Gene2func, gene set analysis, GO molecular functions.

*Sex specific ADHD GWAS:* ADHD GWAS was obtained from sex-specific meta-analyses of case-control ADHD by the Psychiatric Genomics Consortium and the Lundbeck Foundation Initiative for Integrative Psychiatric Research (3). The GWAS included only subjects of European ancestry, 32,102 males (N=14,154 cases & 17,948 controls) and 21,191 females (N=4,945 cases & 16,246 controls), and SNPs with MAF > 0.01 and info-score > 0.8. We excluded duplicated and ambiguous SNPs, which resulted in 5,768,802 SNPs in male-specific GWAS and 5,748,208 SNPs in female-specific GWAS.

**Investigating the interactive effects between polygenic scores for higher fasting insulin and childhood adversity on altered executive function phenotypes in multiple cohorts in a sex-specific manner**

*Participants*

We used data from five prospective birth cohorts: 1) Avon Longitudinal Study of Parents and Children (ALSPAC) (4-6); 2) Maternal Adversity, Vulnerability, and Neurodevelopment (MAVAN) (7); 3) Growing Up in Singapore Towards healthy Outcomes (GUSTO) (8); 4) Adolescent Brain Cognitive Development $^{SM}$ Study (ABCD®); and 5) UK Biobank (UKB) (9) to analyze the gene by environment interaction effects on EF outcomes.

*The Avon Longitudinal Study of Parents and Children (ALSPAC)* (6): The ALSPAC cohort included pregnant women from the county of Avon, UK (4, 5) (N = 14,541) with expected delivery dates between April 1991 and December 1992. Additional recruitment (N = 906) was done later during phases, bringing the total sample size to 15,447. Participants provided informed written consent to participate in the study. Consent for biological samples had been collected in accordance with the Human Tissue Act (2004). Please note that the study website contains details of all the data that is available through a fully searchable data dictionary and variable search tool at http://www.bristol.ac.uk/alspac/researchers/our-data/. For the purpose of our analysis, we included children of 8.5 and 9.5 years old (individuals with peripheral insulin levels data), whose mothers had a pregnancy duration between 37 and 42 weeks, a maternal age at delivery greater than 18 years, a child birthweight greater than 2 kg, child alive at 1 year of age, and we only included singleton pregnancies in the analysis. There were 1,901 males and 1,834 females with complete data available for the analyses.

*Maternal Adversity, Vulnerability, and Neurodevelopment (MAVAN)* Project (7): MAVAN is a birth cohort which follows children from birth up to 6 years of age in Montreal (Quebec) and Hamilton (Ontario), Canada, and has 629 recruited participants (7). Mothers aged 18 years or above, with singleton pregnancies, and fluent in French or English were included in the study. There were 161 subjects with complete data on the predictors used within this study available for the analyses after the exclusion criteria was applied, as described in *supplementary Figure S7*.

*Growing Up in Singapore Towards healthy Outcomes (GUSTO)* (8) prospective cohort: The GUSTO study recruited pregnant women of at least 18 years in age, who were attending their first trimester antenatal ultrasound scan between June 2009 and September 2010 at one of Singapore's two major public maternity units: 1) National University Hospital and 2) KK Women's and Children's Hospital. Initially, the main GUSTO cohort recruited 1,450 mothers. By delivery, from 1216 participants, we excluded twins, subjects without genotyping data available, and participants without complete data for the analysis. Sociodemographic characteristics were collected using standardized self-report questionnaires. Study sample was selected based on data availability for each analysis. There were 466 subjects with complete data on the predictors used within this study available for the analyses, as described in *supplementary Figure S7*.

*Adolescent Brain Cognitive Development* SM Study *(ABCD®)*: This is a large-scale study tracking 9 years old and 10 years old individuals recruited from 21 research sites across the United States. ABCD® Data Release 2.0 which includes 4 waves of data: baseline (N = 11 875), 6-month follow-up (N = 8 623), 12-month follow-up (N = 4 951), and 18-month follow-up (N = 1 919). These data were accessed from the National Institutes of Mental Health Data Archive. There were

7,655 subjects with complete data on the predictors used within this study available for the analyses, as described in *supplementary Figure S7*.

*UK Biobank (UKB)* (9): UKB is a population-based cohort with 502,543 recruited participants between the ages of 37 and 73 from 2006 to 2010 in the UK. This dataset contains information on participants' lifestyle and health data at baseline or follow-up, which were collected through questionnaires, physical measurements, and biological samples. All participants provided informed written consent before data collection. There are a total of 502,543 participants within the cohort. There were 71,036 subjects with complete data on the predictors used within this study available for the analyses, as described in *supplementary Figure S7*.

*Genotyping*

Processing of the genotyping data was done using PLINK 1.9 (10).

*ALSPAC:* Children in ALSPAC cohort were genotyped using the Illumina HumanHap550 quad chip genotyping platform by the Wellcome Trust Sanger Institute, Cambridge, UK and the Laboratory Corporation of America, Burlington, NC, US (11). Standard quality control (QC) procedure was applied: participants with inconsistent self-reported and genotyped sex, minimal or excessive heterozygosity, high levels of individual missingness (>3%) and insufficient sample replication (IBD < 0.8) were excluded. Also, SNPs with call rate < 95%, MAF < 1%, or not in Hardy-Weinberg Equilibrium (HWE) ($p < 5 \times 10^{-7}$) were removed. Following the QC, the genotyping data was imputed using Impute v3 and Haplotype Reference Consortium (HRC) imputation reference panel (release 1.1), which resulted in 38,898,739 SNPs available for analysis. The population structure of ALSPAC cohort was described using principal component analysis (12, 13), which was conducted on the genotyped autosomal SNPs with MAF > 5% with the following pruning parameters for linkage disequilibrium: 100-SNP sliding window, an increment of 5 SNPs, and variance inflation factor (VIF) threshold of 1.01. To account for population stratification, the first ten principal components were included in the analysis.

*MAVAN:* Genome-wide platforms (the Infinium PsychArray v1 or the PsychChip v1.1/v1.2, Illumina, Inc.) were used to genotype 229,456 autosomal SNPs of buccal epithelial cells of children in MAVAN, according to manufacturer's guidelines. SNPs with a low call rate (<95%), low *p*-values on HWE exact test ($p < 1 \times 10^{-30}$), and a MAF < 5% were removed. Afterwards, imputation using the Sanger Imputation Service (McCarthy et al., 2016) and HRC as the reference panel (release 1.1) was performed and SNPs with an info score > 0.8 were retained for the analysis, resulting in 16,249,769 autosomal SNPs. Similar to the ALSPAC cohort, the population structure of the MAVAN cohort was evaluated using principal component analysis of all autosomal SNPs that passed the quality control (MAF > 5%) and not in high linkage disequilibrium ($r^2 > 0.2$) across 50-SNP region and an increment of 5 SNPs (13). Based on the inspection of the scree plot, the first three principal components (PCs) were the most informative of population structure and were included in all subsequent analyses.

*GUSTO*: Genomic DNA was extracted from the frozen umbilical cord specimens for each child and genotyped via Illumina OmniExpress arrays and llumina Exome arrays following the manufacturer's instructions. Similar to the MAVAN cohort, SNPs with a call rate < 95% or a MAF <5% or which deviated from HWE (p < 1e-40) were removed. Samples with call rates < 95% were removed. The Sanger Imputation Service was used to perform genome-wide imputation using 1000G as the reference, which resulted in 4,869,008 SNPs with an info score > 0.8. To assess population structure, we performed principal component analysis on a pruned data set, like the MAVAN cohort. Based on the inspection of the scree plot, the first three principal components

(PCs) were considered as the most informative of population structure in GUSTO cohort and were included in all analyses.

ABCD: The ABCD genetic data on 517,724 genotyped SNPs for 10627 subjects was obtained from ABCD data repository and subjected to the QC procedure, which was carried out using PLINK 1.9 (14). We removed SNPs with a low call rate (<95%), minor allele frequency less than 5% or with low p-values on HWE exact test ($p < 1e\text{-}40$). Also, subjects with a missing call rate > 5% or relatedness issues were excluded. In total, 493,811 autosomal SNPs for 10,329 subjects passed the QC. Then, we imputed the data using the Sanger Imputation Service (15) and the HRC as the reference panel (release 1.1) resulting in 10,027,736 SNPs with an info score > 0.8. The population structure was evaluated using principal component analysis in a similar procedure as for the MAVAN cohort. Frist, from a set of 10,329 subjects we retained 8,873 unrelated subjects by keeping only one subject per family. Then we applied PCA of all autosomal SNPs that passed the quality control (MAF > 5%) and not in high linkage disequilibrium (r2 > 0.2) across 50-SNP region and an increment of 5 SNPs to generate the PCs, then projected the results on the related subjects' subset to obtain the PCs estimates for the remaining subjects. The first ten genetic principal components were included in all subsequent analyses.

*UKB:* Blood samples from UK Biobank were genotyped at the Affymetrix Research Services Laboratory in Santa Clara, California, USA. Genotyping was conducted using a bespoke BiLEVE Axiom array for 50,000 participants and the remaining 450,000 participants were genotyped using the Affymetrix UK Biobank Axiom array. The two SNP arrays are very similar with over 95% common marker content. Axiom Array plates were processed on the Affymetrix GeneTitan® Multi-Channel (MC) Instrument. Genotypes were then called from the resulting intensities in batches of ~4,700 samples (~4,800 including the controls) using the Affymetrix

Power Tools software and the Affymetrix Best Practices Workflow. Individuals with the same genotype at any given SNP will cluster together in a two-dimensional intensity space (one dimension for each targeted allele). For the interim data release, Affymetrix performed further rounds of genotype calling using algorithms customized for the UK Biobank project. These algorithms targeted very rare SNPs with 6 or fewer minor alleles in a batch, and a subset of SNPs for which the generic calling algorithm did not perform optimally. After genotype calling, Affymetrix performed quality control in each batch separately, to exclude SNPs with poor cluster properties. If a SNP did not meet the Affymetrix prescribed QC thresholds in a given batch, it was set to missing in all individuals from that batch. HWE was performed for each batch. Affymetrix also checked sample quality (such as DNA concentration) and genotype calls were provided only for samples with sufficient DNA metrics. For SNP-based QC metrics, only individuals with similar ancestry and the population structure were characterized by computing principal components using only UK Biobank individuals. The array also includes coding variants across a range of minor allele frequencies (MAFs), including rare markers (< 1% MAF); and markers that provide good genome-wide coverage for imputation in European populations in the common (> 5%) and low frequency (1–5%) MAF ranges. More information about the genotyping protocol, QC, and imputation could be found in (16). The population structure of the UK Biobank cohort was evaluated using fastPCA algorithm for principal component analysis (17). To account for population stratification, the first forty principal components were included in the UK Biobank analysis.

*Refined Polygenic Risk Scores (rPRS)*

A polygenic risk score (PRS) is a sum of the genetic effects of many variants, weighted by an estimated effect of association between alleles and the phenotype of interest described in a

GWAS summary statistics. Classically, PRS can be calculated at any p-value thresholds from the GWAS. In this study, we sought to identify the high FI PRS threshold that best predicted FI levels in children from the ALSPAC cohort in a sex-specific manner (using a sex-specific FI GWAS). For that, we used the refined-PRS (rPRS) method previously described in our previous study (18). Subsequently, the rPRS was applied in this study to inspect the interaction effect between the rPRS for higher FI and ELA on EF behaviors in four separate cohorts to assess the effect in four different age groups, separately for males and females.

*ALSPAC:* The FI PRSs were calculated using the FI GWASs separately for males and females ($N_{males}$ = 47,806, $N_{females}$ = 50,404) from the Meta-Analyses of Glucose and Insulin-related traits Consortium (1). Before the PRS calculations, the lists of SNPs from the GWASs were subjected to LD clumping with $r^2$ of 0.2 and ALSPAC cohort as a reference dataset. PRSs were calculated at 100 different p-value thresholds for each individual in the ALSPAC cohort as a sum of the risk alleles count weighted by the effect size described in the GWAS for each SNP (19, 20). Using ALSPAC as a discovery cohort, we utilized Generalized Estimating Equations (GEE) analysis, using R package geepack (21-23), which allows to incorporate several measurements from the same participants to identify the PRS threshold at which the model had the best fit to the data predicting peripheral insulin levels in children at age 8.5-9.5 years, separately in males and females. The best fit model was identified to be with a PRS at $p_{t\text{-intial-males}}$= 0.05 (11,121 SNPs; $N_{males}$=1,901) in males and $p_{t\text{-intial-females}}$=0.15 (27,202 SNPs; $N_{females}$=1,834) in females. To further refine the PRS, a process explained in Batra et al (18) was applied. Precisely, we ran a GEE analysis for each SNP within the identified PRS threshold to find which SNPs were significantly associated with the peripheral insulin levels separately for males and females [$N_{SNP\ males}$= 635 SNPs, $N_{SNP\ females}$ = 1,449 SNPs].

*MAVAN, GUSTO, ABCD, and UKB:* The SNPs we discovered to associate with peripheral insulin levels in the ALSPAC cohort were used to construct a rPRS in these four cohorts. Because the SNPs were selected in a refinement process of a PRS that was created through conventional means, we therefore refer to this PRS as the refined PRS. The rPRS was calculated similarly to the PRS scores in ALSPAC, as a weighted sum of 635 SNPs for males and 1,449 SNPs for females.

*Early Life Adversity*

*MAVAN:* The postnatal adversity score was created by combining the following markers: 1) birth size percentile below 10$^{th}$ percentile or above 90$^{th}$ percentile; 2) gestational age below or equal to 37 weeks; 3) smoking during pregnancy; 4) household total gross income below $30,000; 5) lack of money; 6) presence of domestic violence or sexual abuse during pregnancy; 7) marital strain; 8) pregnancy anxiety; 9) hospitalization in the first 6 months of life; 10) disorganized attachment; 11) poor family function; 12) maternal mental health through the presence of either BDI, EPDS, or STAI. For every item with a continuous score, we used either the 15$^{th}$ or the 85$^{th}$ percentile as the cut-off to add a point to the adversity score. Presence of each component yields one point, and the adversity score represents the summation of the points where the higher the score, the more adversity has been experienced by the individual.

*GUSTO:* The postnatal adversity score was created by combining the following markers: 1) birth size percentile below 10$^{th}$ percentile or above 90$^{th}$ percentile; 2) gestational age below or equal to 37 weeks; 3) smoking during pregnancy; 4) household monthly income below $2000; 5) poor family function; 6) hospitalization in the first 6 months of life; 7) maternal mental health through the presence of either BDI, EPDS, or STAI. For every item with a continuous score, we used either the 15$^{th}$ or the 85$^{th}$ percentile as the cut-off to add a point to the adversity score.

Presence of each component yields one point, and the adversity score represents the summation of the points where the higher the score, the more adversity has been experienced by the individual.

*ABCD:* The postnatal adversity score was created by combining the following markers: 1) birth size below 10$^{th}$ percentile or above 90$^{th}$ percentile; 2) gestational age below or equal to 37 weeks; 3) smoking during pregnancy; 4) being in an incubator after birth; 5) household total gross income below $25,000; 6) presence of verbal, physical, or sexual abuse; 7) youth report of caregiver acceptance; 8) FES conflict; 9) caregiver's mental health. Individuals received 1 point for every item they indicated experiencing and the adversity score represents the summation of the points where the higher the score, the more adversity has been experienced by the individual.

*UKB:* The postnatal adversity score was created by combining the following markers: 1) birth size was below 10$^{th}$ percentile or above 90$^{th}$ percentile (ID20022); 2) maternal smoking (ID1787; 1 point if the answer was yes); 3) feeling hated by family member during childhood (1 point if the answer was sometimes, often, or very often; ID20487); 4) feeling loved during childhood (1 point if the response was never or rarely; ID20489); 5) not having someone to take to the doctor when needed (1 point if the response was often, half a point was given if the response was rarely had someone; ID20491); 6) presence of domestic violence during childhood (physical or sexual) was also counted as one point (ID20488); and 7) whether an individual was part of a multiple birth (1 point if the response was yes; ID1777). Each adverse instrument was given a score of 1 and the adversity score represents the summation of the points where the higher the score, the more adversity has been experienced by the individual.

### Results

#### Sex-specific impulsivity GWAS: Gene-based enrichment analysis

SNPs with $P<10^{-5}$ were selected to describe female and male GWASs. 14 genes were significantly associated with impulsivity in males and 5 in females (*Figures S5 and S6*). 20 genes overlapped between the male and female impulsivity GWASs (out of 101 total genes in males and 114 genes in females) indicating a small degree of shared polygenic architecture between the sexes. Only the female genes exhibited significant association with cellular components, most of which were related to the nervous system: dendritic tree, somatodendritic compartment, neuron projection, synapse, neuron to neuron synapse, and postsynaptic specialization. *ATG13*, which exhibits significant pleiotropy with Alzheimer's disease GWAS and fasting glucose GWAS in addition to reaction time, is enriched in both males and females. Several of the genes enriched in the males GWAS were significantly associated with autism spectrum disorder or schizophrenia GWAS. *MYT1L* and *TSSC1*, genes that are significantly associated with non-planning and motor impulsivity, were enriched only in females.

**Investigating the interactive effects between polygenic scores for higher fasting insulin and childhood adversity on altered executive function phenotypes in multiple cohorts in a sex-specific manner**

*Enrichment Analysis*

Enrichment analyses (MetaCore™) show that the subset of SNPs composing the FI rPRS are significant for several nervous system development processes in a sex-specific manner, as shown in *supplementary Table S1*. Additionally, enrichment analysis (FUMA, https://fuma.ctglab.nl/) of the genes mapped by the SNPs that compose the FI rPRS showed that these genes were significantly associated with certain nervous system development process and GWASs in a sex-specific manner, as shown in *supplementary Table S1.*

**Supplemental Figures and Tables**

Figure S1. Manhattan plot of all the variants analyzed in the UK Biobank for impulsivity in males (N = 128,277).

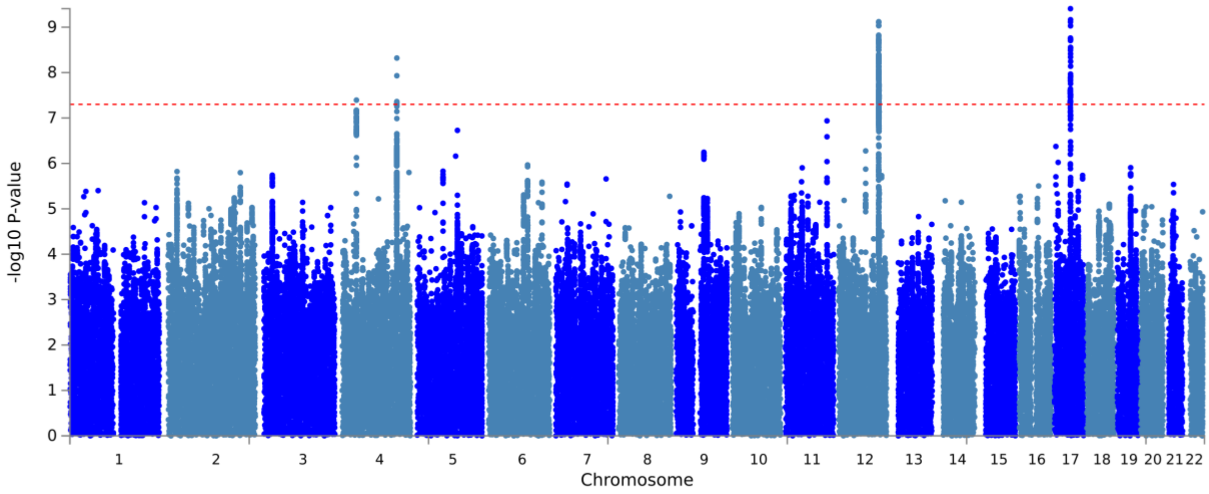


Figure 2. Manhattan plot of all the variants analyzed in the UK Biobank for impulsivity in females (N = 146,607).

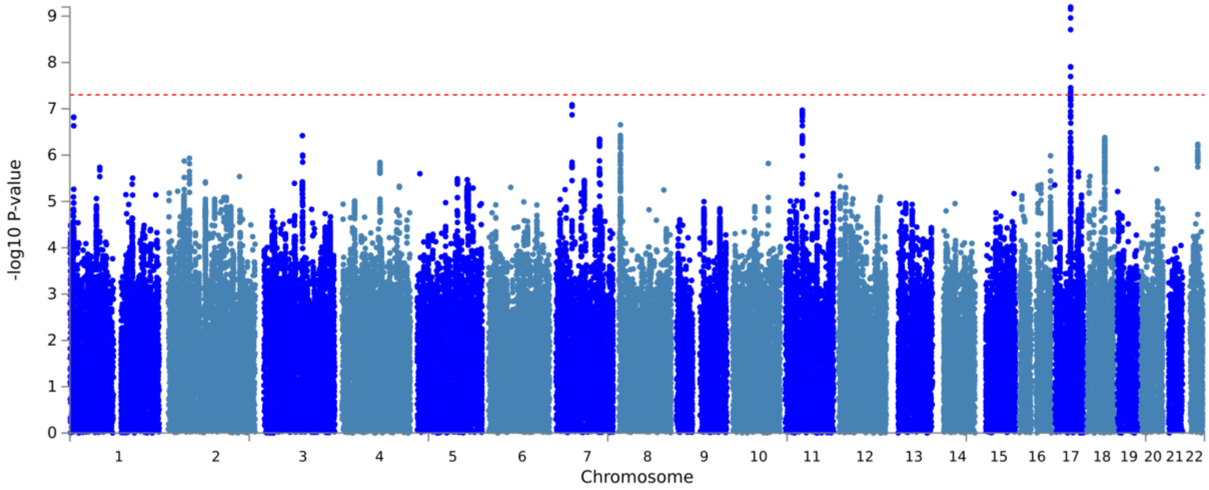

Figure S3. Q-Q plots of the observed P-values on those expected using all the variants analyzed in UK Biobank for impulsivity in males (N = 128,277).

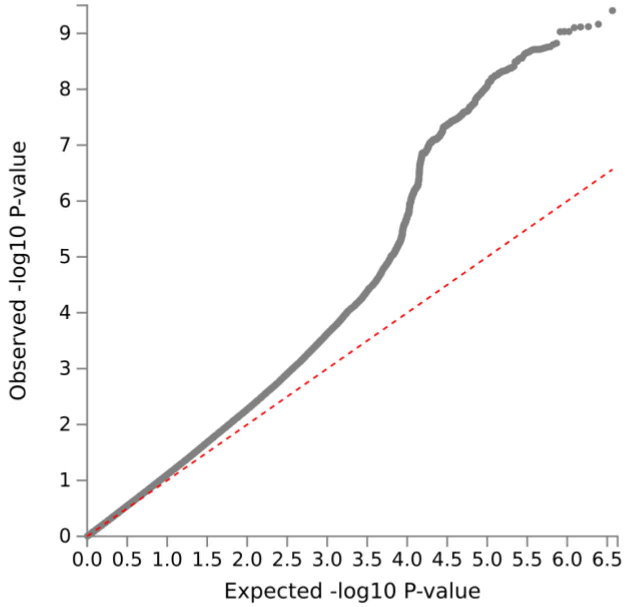


Figure S4. Q-Q plots of the observed P-values on those expected using all the variants analyzed in UK Biobank impulsivity in females (N = 146,607).

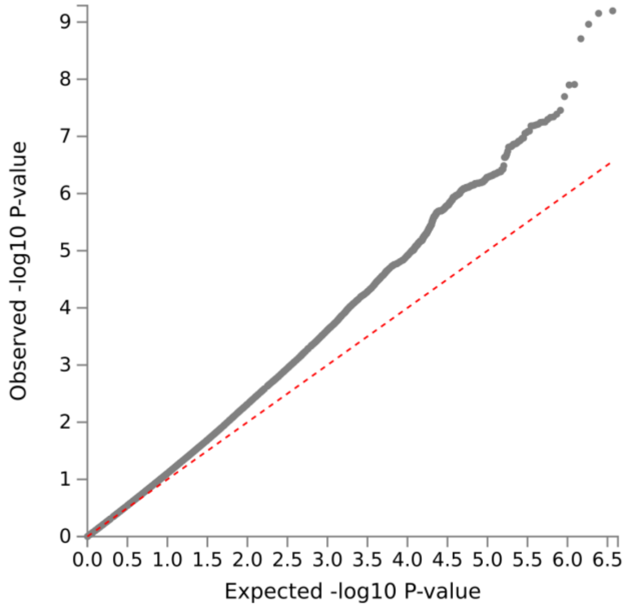

Figure S5. Manhattan plot of the adjusted –log10 P-values of each gene for an association with impulsivity in the UK Biobank cohort in males (N=128,277). The dotted horizontal line represents the gene-based threshold for significance (was defined at P = 0.05/19080 = 2.621e-6, where 19080 is a number of mapped protein coding genes).

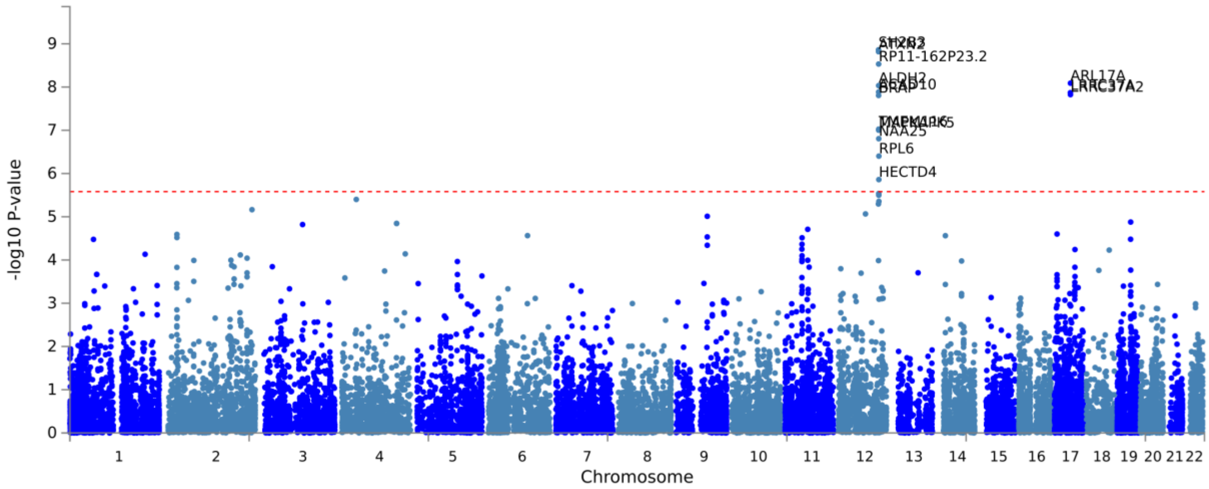


Figure S6. Manhattan plot of the adjusted –log10 P-values of each gene for an association with impulsivity in the UK Biobank cohort in males (N= 146,607). The dotted horizontal line represents the gene-based threshold for significance (was defined at P = 0.05/19080 = 2.621e-6, where 19080 is a number of mapped protein coding genes).

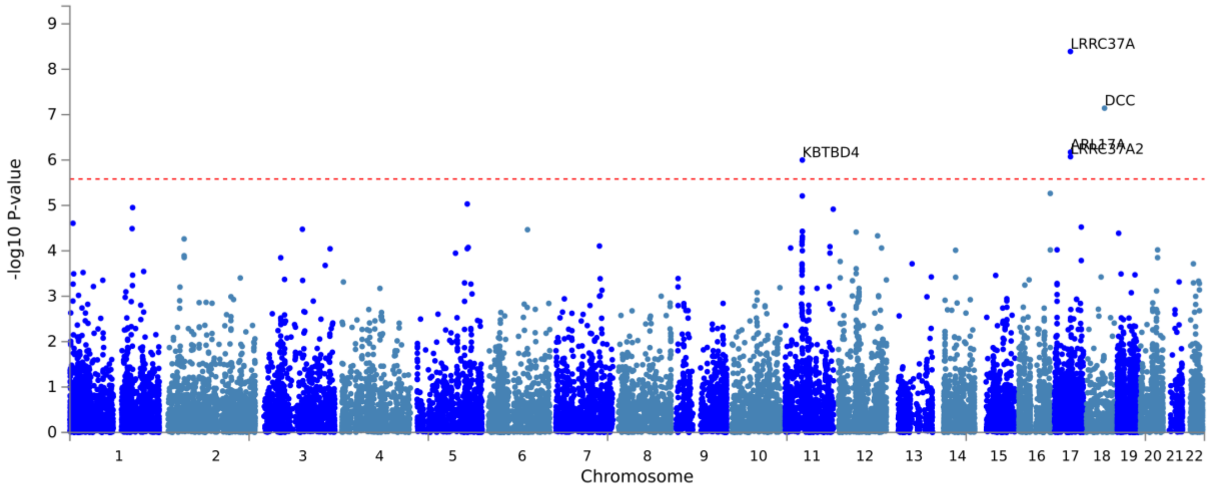

Figure S7. Block scheme describing sample selection in (A) MAVAN, (B) GUSTO, (C) ABCD, and (D) UKB.

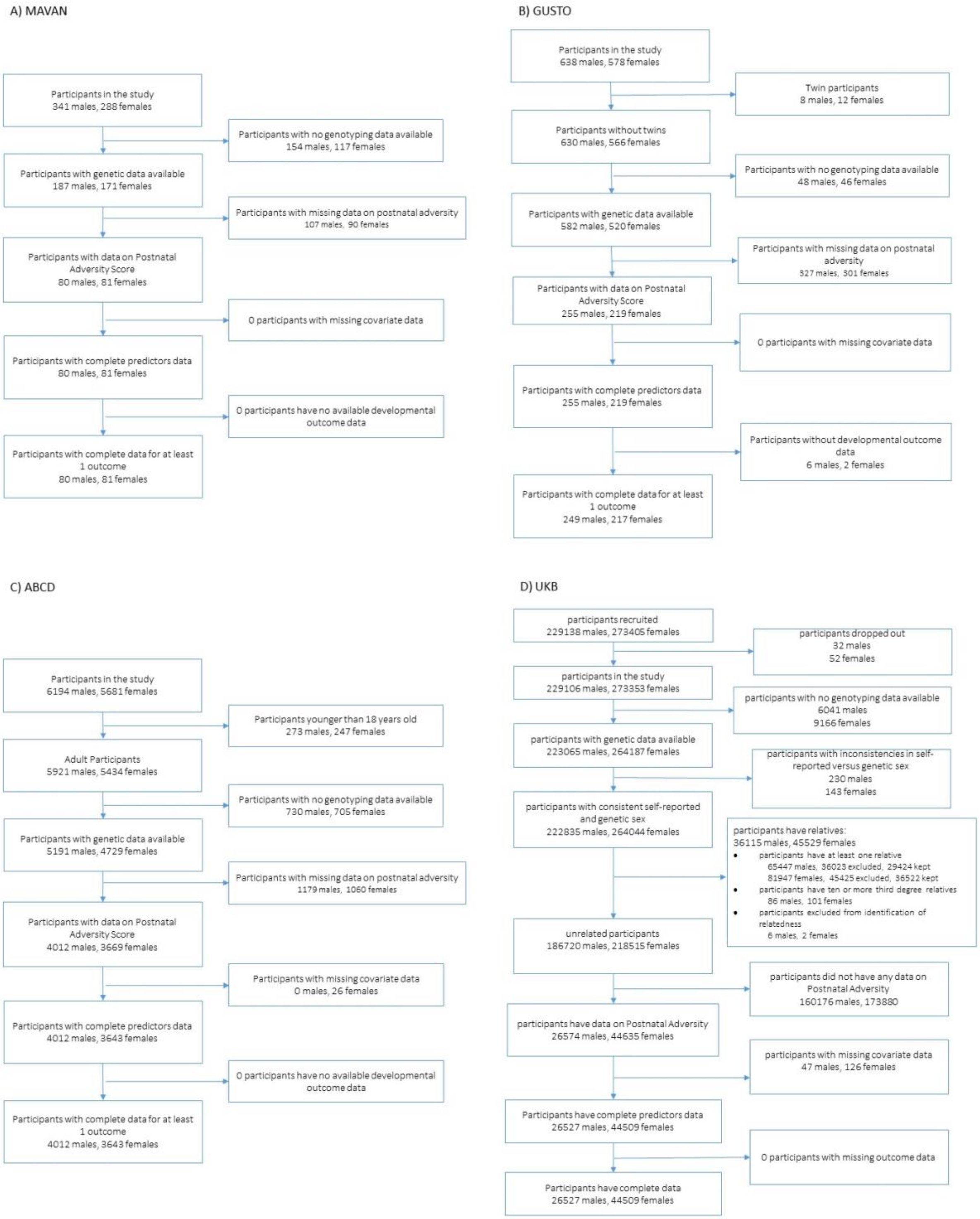

Figure S8. Conditional Q-Q plots (A) fasting insulin conditioned on impulsivity (males), (B) fasting insulin conditioned on impulsivity (females), (C) impulsivity conditioned on fasting insulin (males), (D) impulsivity conditioned on fasting insulin (females), (E) fasting insulin conditioned on ADHD (males), (F) fasting insulin conditioned on ADHD (females), (G) ADHD conditioned on fasting insulin (males), and (H) ADHD conditioned on fasting insulin (females). The plots, (A) and (E) for males and (B) and (F) for females, display the nominal -log10p values of the single SNP association statistics versus their empirical distribution in fasting insulin and below the standard GWAS threshold as a function of significance of association with impulsivity/ADHD at the level of $p < 0.1$, $p < 0.01$, and $p < 0.001$. The plots, (C) and (G) for males and (D) and (H) for females, display the nominal -log10p values of the single SNP association statistics versus their empirical distribution in impulsivity/ADHD and below the standard GWAS threshold as a function of significance of association with fasting insulin at the level of $p < 0.1$, $p < 0.01$, and $p < 0.001$. The blue line indicates all the SNPs. The dashed line indicates the null hypothesis. The blue line indicates all the SNPs. The dashed line indicates the null hypothesis.

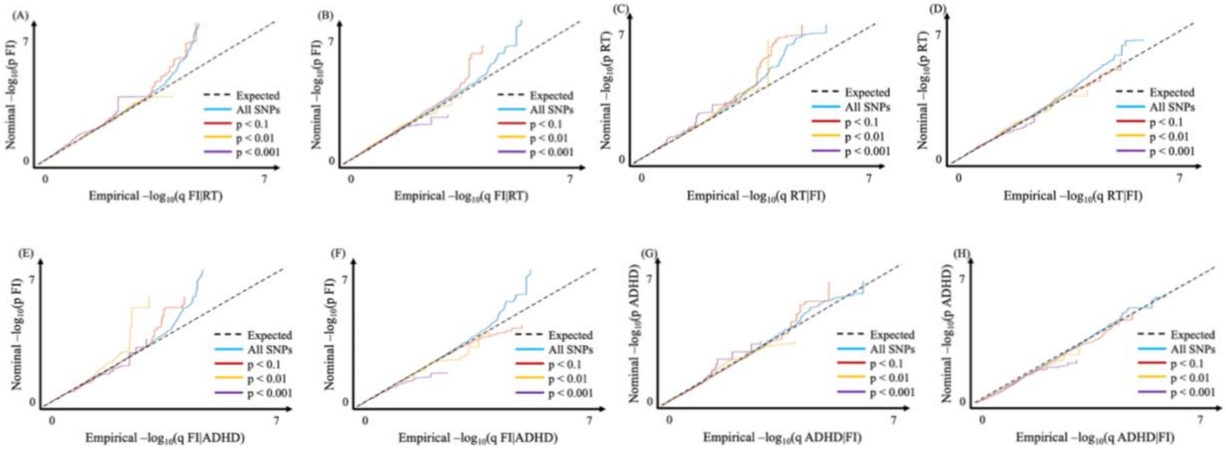

Table S1. Enrichment analyses, through MetaCore™ and FUMA, show that the subset of SNPs and genes composed from the males and females FI rPRS are significant for several nervous system development processes and GWASs.

| **Metacore Results from SNPs composing the FI rPRS** | | |
|---|---|---|
| **Processes** | **Males FDR adjusted p-value** | **Females FDR adjusted p-value** |
| Nervous system disease | $p < 0.001$ | $p < 0.001$ |
| Nervous system development | $p < 0.001$ | $p < 0.001$ |
| Psychiatry and psychology diseases | $p < 0.001$ | |
| Mood disorders | $p < 0.001$ | |
| Neurogenesis axonal guidance | | $p = 0.007$ |
| HTR2A signaling in the nervous system | $p = 0.033$ | $p = 0.019$ |
| Regulation of intrinsic membrane properties and excitability of cortical pyramidal neurons | | $p = 0.0275$ |
| Neurophysiological process dopamine D2 receptor transactivation of PDGFR in the central nervous system | $p = 0.041$ | |
| **FUMA Results from genes mapped from the FI rPRS** | | |
| **GWAS Enrichment** | **Males FDR adjusted p-value** | **Females FDR adjusted p-value** |
| Cognitive ability | $p < 0.001$ | |
| ADHD | $p = 0.0002$ | $p < 0.001$ |
| Cognitive performance | | $p < 0.001$ |
| Food addiction | | $p = 0.0048$ |
| Impulsivity | | $p = 0.018$ |
| **Processes** | **Males FDR adjusted p-value** | **Females FDR adjusted p-value** |
| Biological process cognition | $p = 0.0308$ | $p = 0.00228$ |
| Nervous system disease | $p < 0.001$ | $p < 0.001$ |
| Nervous system development | $p < 0.001$ | $p < 0.001$ |
| Psychiatry and psychology diseases | $p < 0.001$ | |
| Mood disorders | $p < 0.001$ | |
| Neurogenesis axonal guidance | | $p = 0.007$ |
| HTR2A signaling in the nervous system | $p = 0.033$ | $p = 0.019$ |
| Regulation of intrinsic membrane properties and excitability of cortical pyramidal neurons | | $p = 0.0275$ |
| Neurophysiological process dopamine D2 receptor transactivation of PDGFR in the central nervous system | $p = 0.041$ | |

**References**

1. Lagou V, Mägi R, Hottenga J-J, Grallert H, Perry JR, Bouatia-Naji N, et al. (2021): Sex-dimorphic genetic effects and novel loci for fasting glucose and insulin variability. *Nature communications*. 12:1-18.

2. Watanabe K, Taskesen E, van Bochoven A, Posthuma D (2019): FUMA: Functional mapping and annotation of genetic associations. *European Neuropsychopharmacology*. 29:S789-S790.

3. Kettunen J, Demirkan A, Würtz P, Draisma HH, Haller T, Rawal R, et al. (2016): Genome-wide study for circulating metabolites identifies 62 loci and reveals novel systemic effects of LPA. *Nature communications*. 7:1-9.

4. Boyd A, Golding J, Macleod J, Lawlor DA, Fraser A, Henderson J, et al. (2013): Cohort Profile: The 'Children of the 90s'—The Index Offspring of the Avon Longitudinal Study of Parents and Children. *Int J Epidemiol*. 42:111-127.

5. Fraser A, Macdonald-Wallis C, Tilling K, Boyd A, Golding J, Davey Smith G, et al. (2013): Cohort Profile: the Avon Longitudinal Study of Parents and Children: ALSPAC Mothers Cohort. *Int J Epidemiol*. 42:97-110.

6. Northstone K, Lewcock M, Groom A, Boyd A, Macleod J, Timpson N, et al. (2019): The Avon Longitudinal Study of Parents and Children (ALSPAC): An Update on the Enrolled Sample of Index Children in 2019. *Wellcome Open Res*. 4.

7. O'Donnell KA, Gaudreau H, Colalillo S, Steiner M, Atkinson L, Moss E, et al. (2014): The Maternal Adversity, Vulnerability and Neurodevelopment Project: Theory and Methodology. *Can J Psychiatry*. 59:497-508.

8. Soh S-E, Tint MT, Gluckman PD, Godfrey KM, Rifkin-Graboi A, Chan YH, et al. (2014): Cohort profile: Growing Up in Singapore Towards healthy Outcomes (GUSTO) birth cohort study. *International journal of epidemiology*. 43:1401-1409.

9. Sudlow C, Gallacher J, Allen N, Beral V, Burton P, Danesh J, et al. (2015): UK biobank: an open access resource for identifying the causes of a wide range of complex diseases of middle and old age. *PLoS medicine*. 12:e1001779.

10. Chang CC, Chow CC, Tellier LC, Vattikuti S, Purcell SM, Lee JJ (2015): Second-generation PLINK: rising to the challenge of larger and richer datasets. *Gigascience*. 4:s13742-13015-10047-13748.

11. Richmond RC, Timpson NJ, Felix JF, Palmer T, Gaillard R, McMahon G, et al. (2017): Using Genetic Variation to Explore the Causal Effect of Maternal Pregnancy Adiposity on Future Offspring Adiposity: A Mendelian Randomisation Study. *PLoS Med*. 14:e1002221.

12. Patterson N, Price AL, Reich D (2006): Population Structure and Eigenanalysis. *PLoS Genet*. 2:e190.

13. Price AL, Patterson NJ, Plenge RM, Weinblatt ME, Shadick NA, Reich D (2006): Principal Components Analysis Corrects for Stratification in Genome-Wide Association Studies. *Nat Genet*. 38:904-909.

14. Purcell S, Neale B, Todd-Brown K, Thomas L, Ferreira MA, Bender D, et al. (2007): PLINK: a tool set for whole-genome association and population-based linkage analyses. *The American journal of human genetics*. 81:559-575.

15. McCarthy S, et al. (2016): A reference panel of 64,976 haplotypes for genotype imputation. *Nature genetics*. 48:1279-1283.

16. Bycroft C, Freeman C, Petkova D, Band G, Elliott LT, Sharp K, et al. (2018): The UK Biobank resource with deep phenotyping and genomic data. *Nature*. 562:203.

17. Galinsky KJ, Bhatia G, Loh P-R, Georgiev S, Mukherjee S, Patterson NJ, et al. (2016): Fast principal-component analysis reveals convergent evolution of ADH1B in Europe and East Asia. *The American Journal of Human Genetics*. 98:456-472.

18. Batra A, Chen LM, Wang Z, Parent C, Pokhvisneva I, Patel S, et al. (2021): Early life adversity and polygenic risk for high fasting insulin are associated with childhood impulsivity. *Frontiers in neuroscience*. 15.

19. Wray NR, Lee SH, Mehta D, Vinkhuyzen AA, Dudbridge F, Middeldorp CM (2014): Research Review: Polygenic Methods and Their Application to Psychiatric Traits. *J Child Psychol Psychiatry*. 55:1068-1087.

20. Dudbridge F (2013): Power and Predictive Accuracy of Polygenic Risk Scores. *PLoS Genet*. 9.

21. Højsgaard S, Halekoh U, Yan J (2006): The R package geepack for generalized estimating equations. *Journal of statistical software*. 15:1-11.

22. Yan J, Fine J (2004): Estimating equations for association structures. *Statistics in medicine*. 23:859-874.

23. Yan J (2002): Geepack: yet another package for generalized estimating equations. *R-news*. 2:12-14.
